# East Asian-specific and cross-ancestry genome-wide meta-analyses provide mechanistic insights into peptic ulcer disease

**DOI:** 10.1101/2022.10.25.22281344

**Authors:** Yunye He, Masaru Koido, Yoichi Sutoh, Mingyang Shi, Yayoi Otsuka-Yamasaki, Hans Markus Munter, Takayuki Morisaki, Akiko Nagai, Yoshinori Murakami, Chizu Tanikawa, Tsuyoshi Hachiya, Koichi Matsuda, Atsushi Shimizu, Yoichiro Kamatani

## Abstract

Peptic ulcer disease (PUD) refers to acid-induced injury of the digestive tract, occurring mainly in the stomach (gastric ulcer; GU) or duodenum (duodenal ulcer; DU). We conducted a large-scale cross-ancestry meta-analysis of PUD combining genome-wide association studies with four Japanese and two European studies (52,032 cases and 905,344 controls), and discovered 25 novel loci highly concordant across ancestries. Based on these loci, an examination of similarities and differences in genetic architecture between GU and DU demonstrated that GU shared the same risk loci as DU, although with smaller genetic effect sizes and higher polygenicity than DU, indicating higher heterogeneity of GU. *H. pylori* (HP)-stratified analysis found an HP-related host genetic locus, marking its role in HP-mediated PUD etiology. Integrative analyses using bulk and single-cell transcriptome profiles highlighted the genetic factors of PUD to be enriched in the highly expressed genes in stomach tissues, especially in somatostatin-producing D cells. Our results provide genetic evidence that gastrointestinal cell differentiations and hormone regulations are critical in PUD etiology.

---

Peptic ulcer disease (PUD) refers to the acid-induced injury of digestive tract, occurring mainly in the stomach (gastric ulcer; GU) or proximal segment of the duodenum (duodenal ulcer; DU) with bleeding, perforation, or gastric outlet obstruction as the major complication. PUD is one of the most common gastrointestinal disorders, with a lifetime prevalence rate of approximately 5–10% in the general population^1^. The prevalence of PUD has been reported to be substantially higher in East Asians (EAS) than in Europeans (EUR)^1^, with GU being more common than DU in the Japanese population and DU being more common in Europeans^2^.

With *H. pylori* (HP) infection and the use of non-steroidal anti-inflammatory drugs (NSAIDs) being two of the most common causes of GU and DU^3^, genetic factors also play a critical role in the development of PUD^4^. Previous genome-wide association studies (GWASs) of PUD had identified multiple loci, mainly HP-related, in Europeans^5,6^. Given the relatively high prevalence of PUD and HP infection in East Asians and the remarkably limited number of risk loci identified in East Asian populations^2,7^, GWAS with a larger sample size of EAS ancestry individuals would be required to enhance our understanding of genetic etiology of PUD. Since GU and DU differ in various aspects, such as the proportion of ulcers that are attributable to HP infection^8^, the genetic differences across PUD subtypes and the key cell types involved in their etiology should be investigated. Epidemiological studies have suggested that DU is a protective factor against gastric cancer (GC)^9^. However, whether genetic factors for PUD and GC are concordant and can explain the epidemiological findings remains unclear. Therefore, we anticipated that large-scale genetic studies of PUD could not only expand our understanding of PUD biology but also provide insights into the genetic factors that interact with *H. pylori* or lead to different outcomes of PUD or GC, potentially enabling a more accurate prediction of individual risk in clinical settings.

To address these issues, we conducted the largest-to-date East Asian-specific and cross-ancestry genome-wide analysis of PUD and PUD subtypes along with four Japanese studies and two European cohorts totaling 52,032 PUD cases and 905,344 controls.

## Results

We conducted a three-stage genome-wide analysis of PUD and its subtypes. An overview of the workflow is provided in **Fig. 1** and **Supplementary Figure 1**. PUD cases in the East Asian populations were obtained by combining individuals with any of the two major PUD subtypes (DU and GU), which were classified based on the anatomical sites where peptic ulcers occurred. Individuals with comorbidities of GU and DU were classified as BU (both GU and DU) cases (**Methods; Supplementary Figure 2; Supplementary Table 1**).

**Fig. 1.**
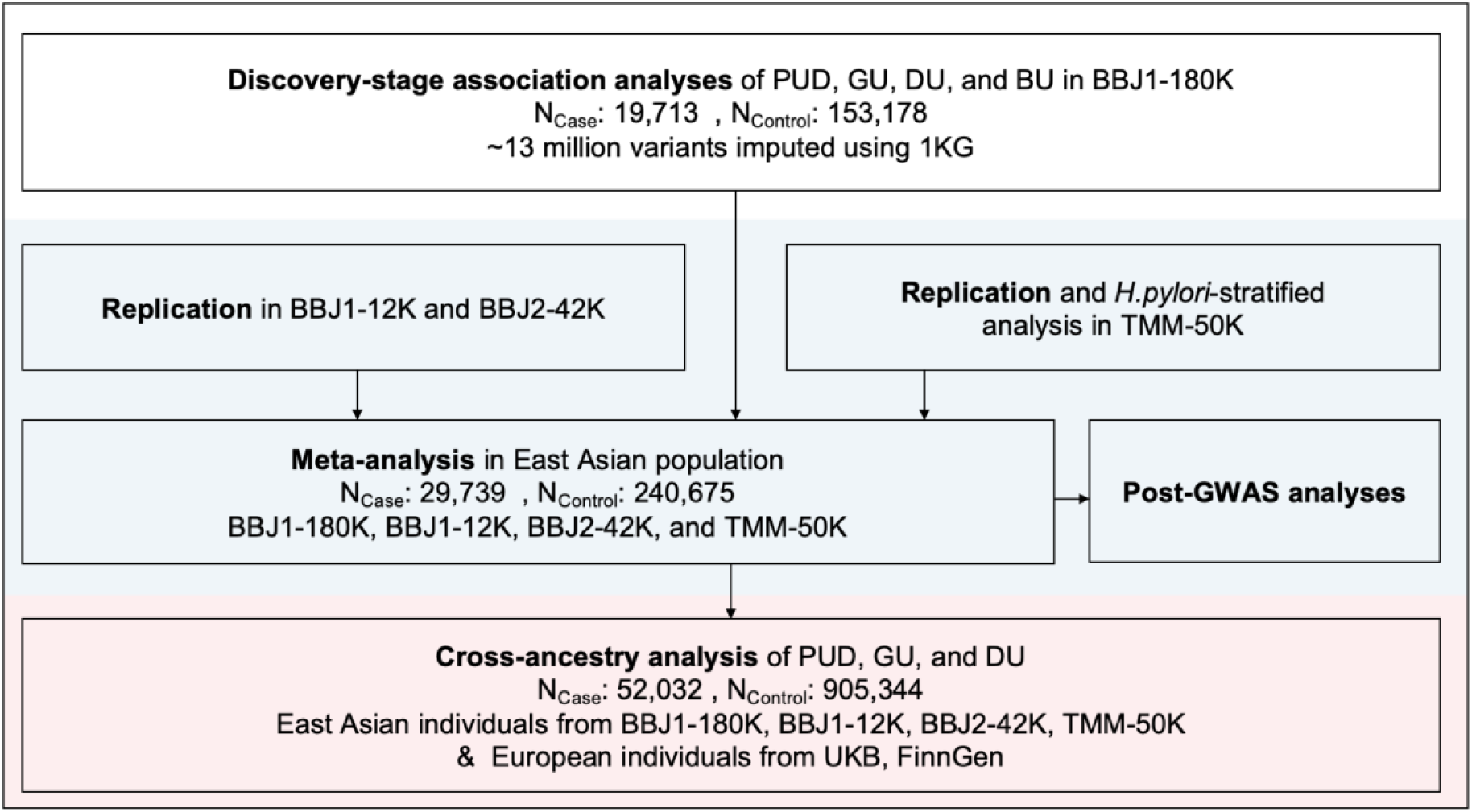
Overview of the three-stage study design. 1KG, 1000 Genomes Project panel; PUD, peptic ulcer diseases; GU, gastric ulcers; DU, duodenal ulcers; BU, comorbidities of DU and GU; BBJ1-180K, approximately 180K individuals from Biobank Japan 1^st^ cohort; BBJ1-12K, approximately 12K individuals from BioBank Japan 1^st^ cohort; BBJ2-42K, approximately 42K individuals from Biobank Japan 2^nd^ cohort; TMM-50K, approximately 50K individuals from Tohoku Medical Megabank; UKB, UK Biobank

### Association analyses of PUD and its subtypes

First, we performed GWAS of PUD and PUD subtypes (DU, GU, and BU) in the discovery stage on the BioBank Japan first cohort (BBJ1)^10^-180K dataset (**Methods**). The dataset included 19,713 PUD cases and 153,178 controls of East Asian ancestry and was imputed using 1000 Genomes Project phase 3^11^ (1KG Phase 3) reference panel. A total of 13,846,852 variants (minor allele count > 20 and Rsq > 0.3) were tested for association with a generalized linear mixed model using SAIGE^12^, which controls for the case-control imbalance (case-to-control ratios ranged from 1:7.7 to 1:82 in BBJ1-180K; **Supplementary Table 1**). For PUD, ten genome-wide significant loci (*P* < 5.0 × 10^−8^) were identified, five of which had not been reported as genome-wide significant loci in previous GWASs of PUD or any subtype. Additionally, 14 loci reached the significance threshold for DU, including seven novel loci (three of which overlapped with novel PUD loci). One previously reported locus at *PSCA*^*2*^ was identified for GU and BU. A total of 15 non-overlapping genetic loci (**Methods**) reached the significance threshold for PUD or any subtype, of which 9 were novel (**Supplementary Table 2; Supplementary Figure 3**). Analysis of the X chromosome identified one locus at *GUCY2F*^7^ reported for PUD, DU, and GU (**Supplementary Table 3**). Thirteen non-overlapping significant loci were identified in sex-stratified analysis (13 for males and 1 for females; **Supplementary Table 3**).

Replication was conducted in individuals from three independent studies, namely BBJ1-12K (1,001 cases), BBJ2-40K (3,637 cases), and TMM^13^-50K (a population-based study; 5,388 cases; **Supplementary Table 1; Methods)**. The replication datasets were imputed using 1KG Phase 3 panel and tested for associations of autosomal variants with the same settings as in discovery GWAS. Among the nine novel lead variants associated with PUD or subtypes, four were nominally associated (*P* < 0.05) with PUD or its subtypes in the same direction in at least two replication datasets. Notably, five novel loci were replicated in the population-based dataset (*P* < 0.05 in the same direction; **Supplementary Table 4**).

Next, we performed an East Asian-specific meta-analysis combining the discovery GWAS and three replication GWAS (N_case_ = 29,739; N_control_ = 240,675). Fixed-effect meta-analyses using the inverse-variance-weighted (IVW) method were performed for PUD and PUD subtypes. The genomic inflation factors (λ_gc_) and linkage disequilibrium (LD) score regression (LDSC)^14^ intercepts ranged from 1.03 to 1.08 and from 1.01 to 1.02, respectively (**Supplementary Table 5)**, indicating no substantial bias. In the EAS-specific meta-analysis, we detected 25 non-overlapping risk loci associated with PUD or any subtype, including 11 additional novel loci (**Table 1**; **Supplementary Figure 4; Supplementary Table 6**).

**Table 1.**
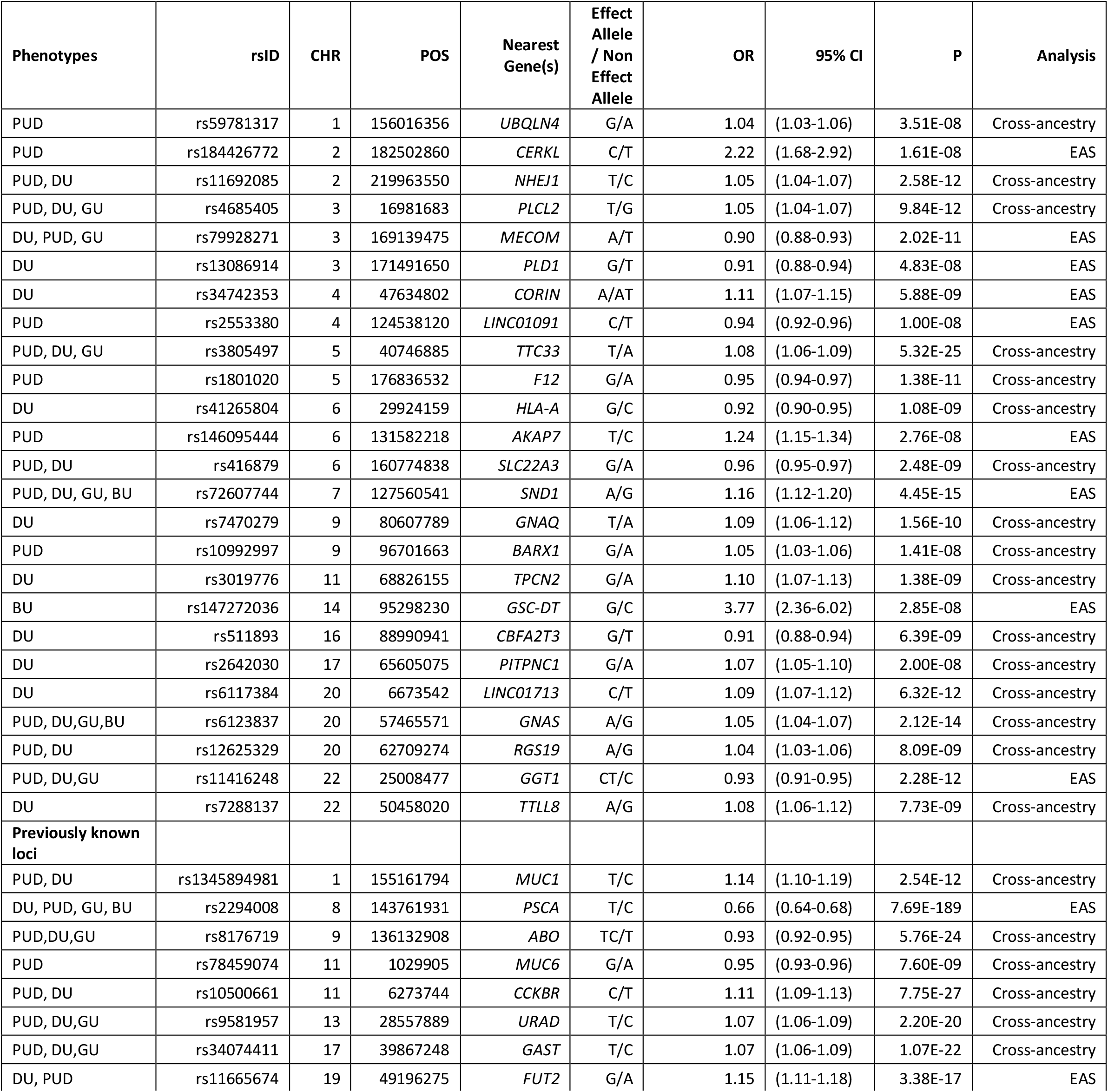
Significant loci associated with PUD or PUD subtypes from genome-wide meta-analyses. Lead variants from each significant locus identified in fixed-effect population-specific or cross-ancestry meta-analysis are shown. Only the most significant lead variant is shown for loci associated with multiple phenotypes or identified in multiple analyses. Variants were annotated with the nearest genes. Genomic coordinates were mapped against GRCh37 (hg19). The analysis that first identified the locus is shown.

| Phenotypes | rsID | CHR | POS | Nearest Gene(s) | Effect Allele / Non Effect Allele | OR | 95% CI | P | Analysis |
| --- | --- | --- | --- | --- | --- | --- | --- | --- | --- |
| PUD | rs59781317 | 1 | 156016356 | UBQLN4 | G/A | 1.04 | (1.03-1.06) | 3.51E-08 | Cross-ancestry |
| PUD | rs184426772 | 2 | 182502860 | CERKL | C/T | 2.22 | (1.68-2.92) | 1.61E-08 | EAS |
| PUD, DU | rs11692085 | 2 | 219963550 | NHEJ1 | T/C | 1.05 | (1.04-1.07) | 2.58E-12 | Cross-ancestry |
| PUD, DU, GU | rs4685405 | 3 | 16981683 | PLCL2 | T/G | 1.05 | (1.04-1.07) | 9.84E-12 | Cross-ancestry |
| DU, PUD, GU | rs79928271 | 3 | 169139475 | MECOM | A/T | 0.90 | (0.88-0.93) | 2.02E-11 | EAS |
| DU | rs13086914 | 3 | 171491650 | PLD1 | G/T | 0.91 | (0.88-0.94) | 4.83E-08 | EAS |
| DU | rs34742353 | 4 | 47634802 | CORIN | A/AT | 1.11 | (1.07-1.15) | 5.88E-09 | EAS |
| PUD | rs2553380 | 4 | 124538120 | LINC01091 | C/T | 0.94 | (0.92-0.96) | 1.00E-08 | EAS |
| PUD, DU, GU | rs3805497 | 5 | 40746885 | TTC33 | T/A | 1.08 | (1.06-1.09) | 5.32E-25 | Cross-ancestry |
| PUD | rs1801020 | 5 | 176836532 | F12 | G/A | 0.95 | (0.94-0.97) | 1.38E-11 | Cross-ancestry |
| DU | rs41265804 | 6 | 29924159 | HLA-A | G/C | 0.92 | (0.90-0.95) | 1.08E-09 | Cross-ancestry |
| PUD | rs146095444 | 6 | 131582218 | AKAP7 | T/C | 1.24 | (1.15-1.34) | 2.76E-08 | EAS |
| PUD, DU | rs416879 | 6 | 160774838 | SLC22A3 | G/A | 0.96 | (0.95-0.97) | 2.48E-09 | Cross-ancestry |
| PUD, DU, GU, BU | rs72607744 | 7 | 127560541 | SND1 | A/G | 1.16 | (1.12-1.20) | 4.45E-15 | EAS |
| DU | rs7470279 | 9 | 80607789 | GNAQ | T/A | 1.09 | (1.06-1.12) | 1.56E-10 | Cross-ancestry |
| PUD | rs10992997 | 9 | 96701663 | BARX1 | G/A | 1.05 | (1.03-1.06) | 1.41E-08 | Cross-ancestry |
| DU | rs3019776 | 11 | 68826155 | TPCN2 | G/A | 1.10 | (1.07-1.13) | 1.38E-09 | Cross-ancestry |
| BU | rs147272036 | 14 | 95298230 | GSC-DT | G/C | 3.77 | (2.36-6.02) | 2.85E-08 | EAS |
| DU | rs511893 | 16 | 88990941 | CBFA2T3 | G/T | 0.91 | (0.88-0.94) | 6.39E-09 | Cross-ancestry |
| DU | rs2642030 | 17 | 65605075 | PITPNC1 | G/A | 1.07 | (1.05-1.10) | 2.00E-08 | Cross-ancestry |
| DU | rs6117384 | 20 | 6673542 | LINC01713 | C/T | 1.09 | (1.07-1.12) | 6.32E-12 | Cross-ancestry |
| PUD, DU, GU, BU | rs6123837 | 20 | 57465571 | GNAS | A/G | 1.05 | (1.04-1.07) | 2.12E-14 | Cross-ancestry |
| PUD, DU | rs12625329 | 20 | 62709274 | RGS19 | A/G | 1.04 | (1.03-1.06) | 8.09E-09 | Cross-ancestry |
| PUD, DU, GU | rs11416248 | 22 | 25008477 | GGT1 | CT/C | 0.93 | (0.91-0.95) | 2.28E-12 | EAS |
| DU | rs7288137 | 22 | 50458020 | TTLL8 | A/G | 1.08 | (1.06-1.12) | 7.73E-09 | Cross-ancestry |
| Previously known loci |  |  |  |  |  |  |  |  |  |
| PUD, DU | rs1345894981 | 1 | 155161794 | MUC1 | T/C | 1.14 | (1.10-1.19) | 2.54E-12 | Cross-ancestry |
| DU, PUD, GU, BU | rs2294008 | 8 | 143761931 | PSCA | T/C | 0.66 | (0.64-0.68) | 7.69E-189 | EAS |
| PUD, DU, GU | rs8176719 | 9 | 136132908 | ABO | TC/T | 0.93 | (0.92-0.95) | 5.76E-24 | Cross-ancestry |
| PUD | rs78459074 | 11 | 1029905 | MUC6 | G/A | 0.95 | (0.93-0.96) | 7.60E-09 | Cross-ancestry |
| PUD, DU | rs10500661 | 11 | 6273744 | CCKBR | C/T | 1.11 | (1.09-1.13) | 7.75E-27 | Cross-ancestry |
| PUD, DU, GU | rs9581957 | 13 | 28557889 | URAD | T/C | 1.07 | (1.06-1.09) | 2.20E-20 | Cross-ancestry |
| PUD, DU, GU | rs34074411 | 17 | 39867248 | GAST | T/C | 1.07 | (1.06-1.09) | 1.07E-22 | Cross-ancestry |
| DU, PUD | rs11665674 | 19 | 49196275 | FUT2 | G/A | 1.15 | (1.11-1.18) | 3.38E-17 | EAS |

Finally, we collected publicly available European GWASs of PUD and its subtypes using samples from FinnGen and UK Biobank^5,12,15^ (**Supplementary Table 7**). After QC and harmonization (**Methods**), a fixed-effect IVW cross-ancestry meta-analysis (52,032 PUD cases and 905,344 controls) was performed, combining the Japanese and European studies. Six additional loci for PUD and DU reached the genome-wide significance level (*P* < 5.0 × 10^−8^; **Table 1; Fig. 2; Supplementary Table 8; Supplementary Figure 5**). Further, we performed a cross-ancestry meta-regression utilizing MR-MEGA^16^, and identified 23 known and described novel loci mentioned above in the East Asian-specific and cross-ancestry meta-analysis (**Supplementary Table 9**). In total, we identified 25 non-overlapping novel loci for PUD and its subtypes in the East Asian-specific and cross-ancestry meta-analyses, although two novel loci identified in the discovery stage were not significant in any of the meta-analyses (**Supplementary Table 2**).

**Fig. 2.**
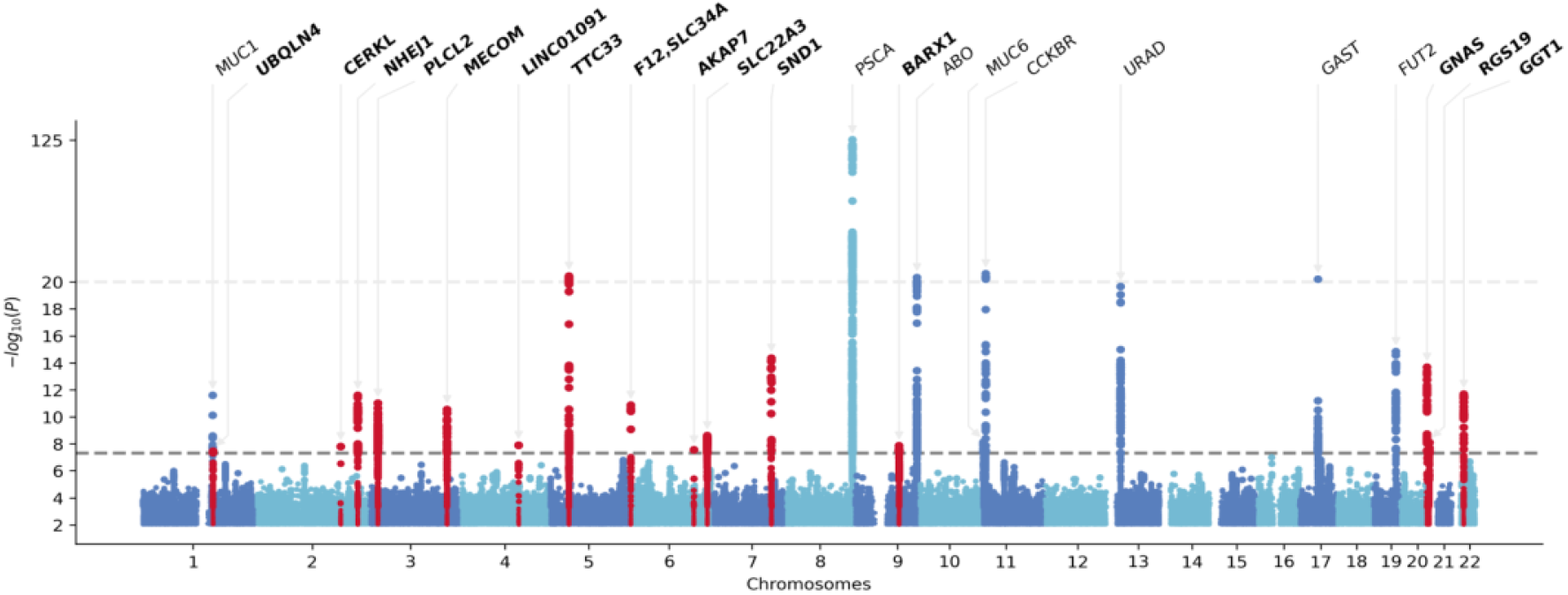
Manhattan plot of the cross-ancestry meta-analysis for peptic ulcer diseases. P values were derived from the cross-ancestry meta-analysis of 52,032 cases and 905,344 controls of EAS or EUR ancestry. Meta-analysis was performed using the inverse-variance weighted method under the fixed-effect model. For variants above the top light grey dashed line (–log_10_(P) >20), values are rescaled. Lead variants are annotated with the nearest gene name. Novel loci are highlighted in red. Variants are plotted against GRCh37 (hg19). The bottom dark grey dashed line indicates the genome-wide significance threshold (*P* < 5.0 × 10^−8^). Variants with –log_10_(P) < 2 were omitted.

### Cross-ancestry effect size and genetic correlation comparison

With the largest available datasets for PUD and its subtypes in EAS to date, we investigated the shared and distinct risk loci for PUD in EAS and EUR individuals. We compared the per-allele effect sizes of lead variants associated with PUD or any of the subtypes available for both ancestries (**Methods**). The effect sizes for PUD showed a relatively high correlation (27 variants with MAF > 0.01 in both populations; r^2^ = 0.76) between the two ancestries, although we detected 10 variants (10/27 = 37%) with significant heterogeneity (P_het_ < 0.05), and only five variants near *PSCA, LINC01091, HIST1H2BD, MUC1*, and *MUC6* loci showed relatively large effect size differences (difference in log(OR) > 0.05; **Fig. 3a; Supplementary Table 10**). To further examine the difference in genetic architecture of PUD between East Asians and Europeans, we conducted a cross-ancestry genetic correlation analysis using Popcorn^17^ (**Methods**). The genetic impact was significantly different from one (null hypothesis: Pgi = 1) for PUD (**Fig. 3b**, genetic impact correlation P_gi_ = 0.65, P = 3.0 × 10^−4^), indicating the difference in genetic architecture of PUD across ancestries. For the subtypes, effect sizes for DU showed a higher correlation (r^2^ = 0.79) across ancestries compared to that for GU (r^2^ = 0.63**; Supplementary Table 11; Supplementary Figure 6**). It was noteworthy that the genetic correlation of GU was relatively low (P_gi_ = 0.45, P = 7.3 × 10^−3^), whereas the genetic architecture of DU did not show a significant difference across ancestries (P_gi_ = 0.72, P = 9.6 × 10^−2^; **Supplementary Table 11**).

**Fig. 3.**
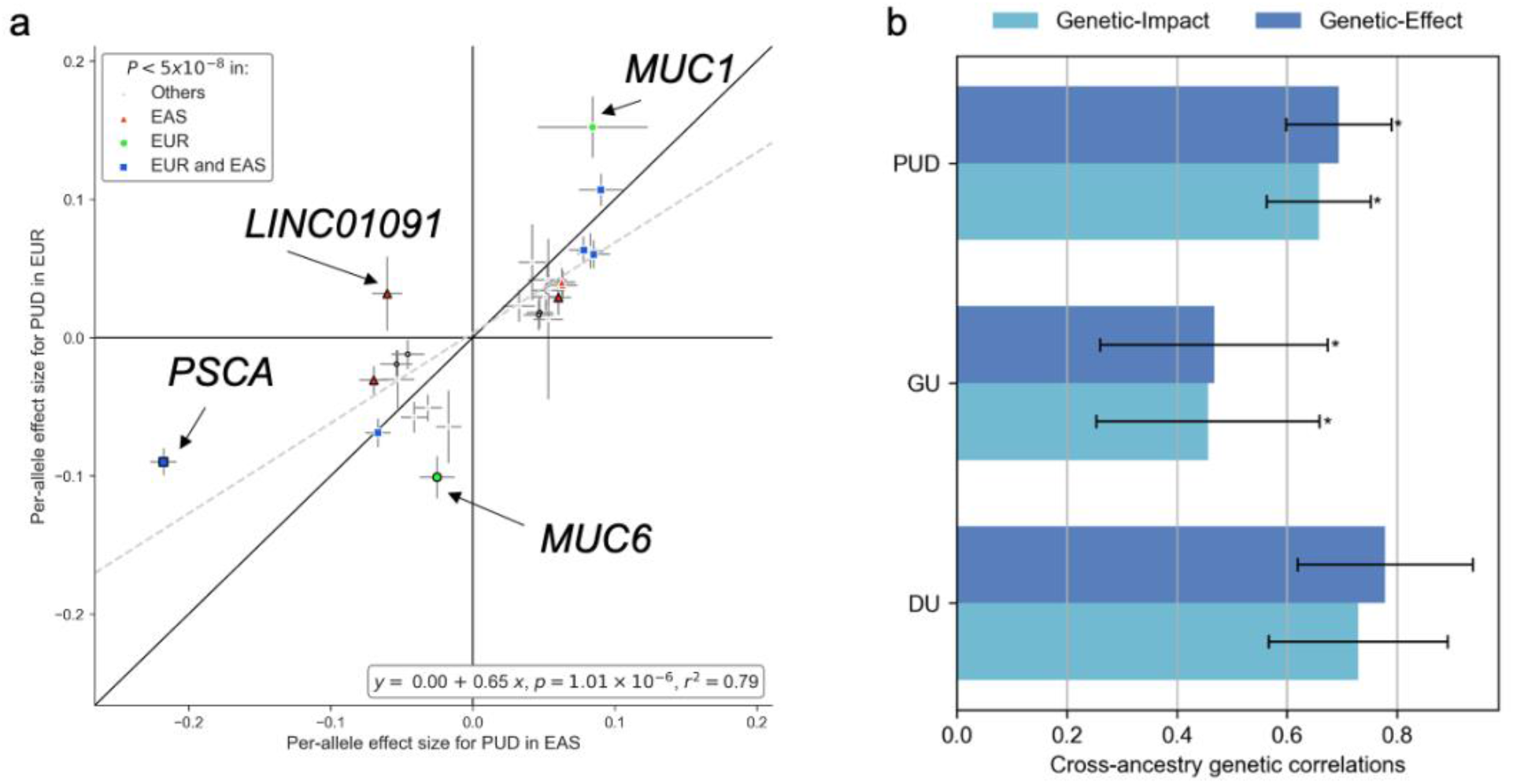
Cross-ancestry effect size comparison and genetic correlation analysis. **a**. Per-allele effect size (logarithm of odds ratios) comparison using East Asian-specific and European-specific summary statistics for PUD. Lead variants associated with PUD or any subtype in East Asian-specific, European-specific, or cross-ancestry meta-analysis were selected for comparison. The most significant associations were shown if overlapping variants existed (interval < 500 kb). Variants with nominally significant heterogeneity (P_het_ < 0.05) were denoted by the black marker edges. The grey dashed line represents the fitted linear regression line. **b**. Cross-ancestry genetic correlation for PUD, GU, and DU estimated by Popcorn. Asterisks indicate estimates that were significantly less than one after Bonferroni correction (P < 0.05/6). Error bars represent the standard error.

### Characterization of PUD-associated loci in East Asians

To explore the secondary signals at the identified loci, we conducted a stepwise conditional analysis using COJO^18^ with an in-sample LD reference for EAS (**Methods**). We detected four additional independent signals reaching genome-wide significance (*P* < 5.0 × 10^−8^) for PUD and three independent signals at *PSCA* locus for DU (**Supplementary Table 12**). *PSCA* locus had the largest number of independent associations (three for PUD, four for DU, and two for GU and BU). Near *CDX2* and *GAST* loci, two of the previously reported loci in European individuals^5^, we detected independent signals at *PDX1* (**Fig. 4a**) and *JUP2* (**Fig. 4b**) loci, respectively.

**Fig. 4.**
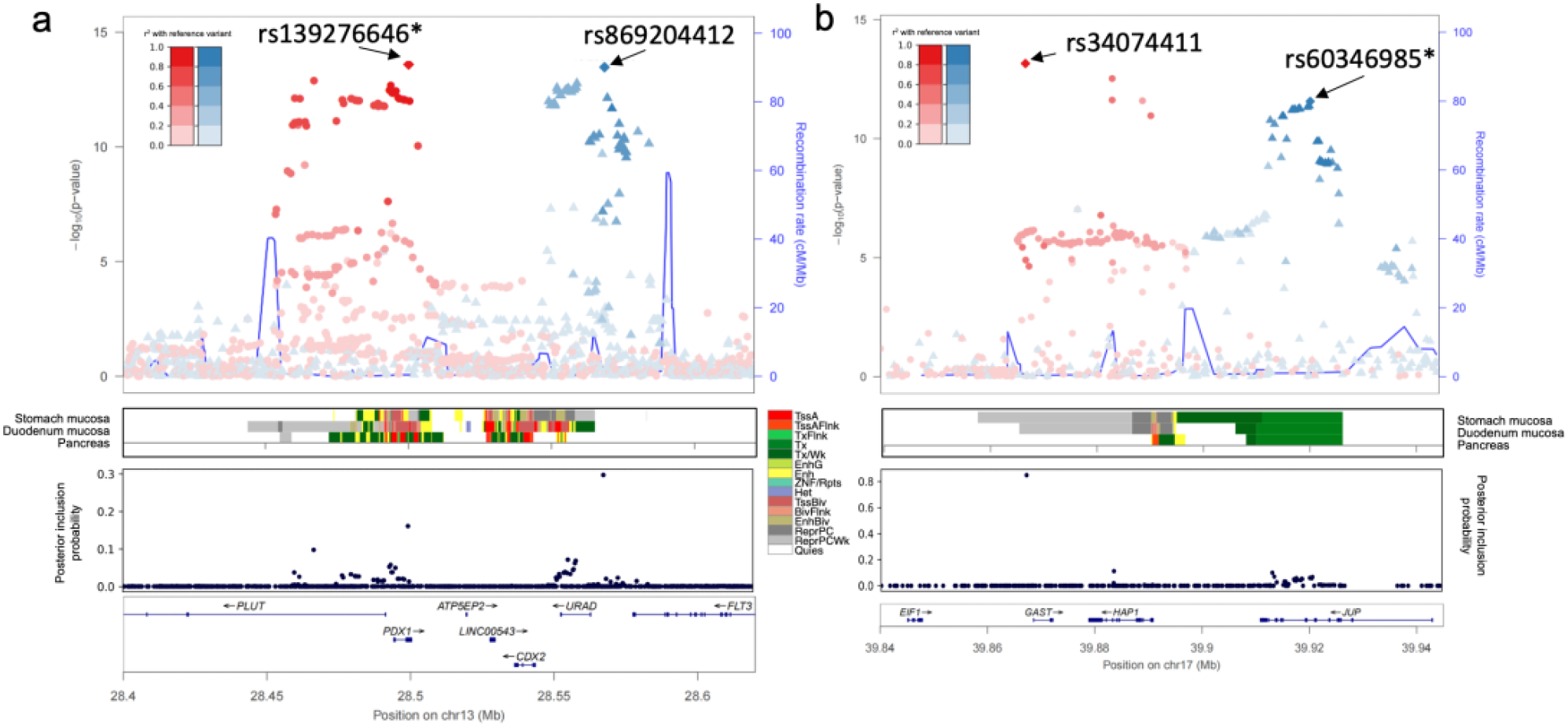
EAS-specific secondary signals at PDX1 and JUP. **a**. Regional plot at *PDX1-CDX2* locus for PUD association from East Asian-specific meta-analysis. Variants are colored to match the lead SNPs in the highest LD, and the extent of LD with the lead variant is shown by a color gradient (red or blue). Posterior inclusion probability (PIP) was derived from fine-mapping analysis. Variants are plotted against GRCh37 (hg19). Chromatin states (core 15-state model) are shown for three related tissue types, namely stomach mucosa, duodenal mucosa, and pancreas. TssA, active transcription start site (TSS); TssAFlnk, flanking active TSS; TxFlnk, transcription at gene 5′ and 3′ ends; Tx, strong transcription; TxWk, weak transcription; EnhG, genic enhancers; Enh, enhancers; ZNF/Rpts, ZNF genes & repeats; Het, heterochromatin; TssBiv, bivalent/poised TSS; BivFlnk, flanking bivalent TSS/Enh; EnhBiv, bivalent enhancer; ReprPC, repressed PolyComb; ReprPCWk, weak repressed PolyComb; Quies, quiescent/low. **b**. Regional plot at *GAST-JUP* locus for PUD association from East Asian-specific meta-analysis.

We conducted a fine-mapping analysis using SuSiE^19^ to identify the causal variants. We searched for nonsynonymous variants in 95% credible sets to link the disease-associated loci to potential alteration of protein functions. A total of 10 nonsynonymous variants at six non-overlapping loci were identified, six of which were in novel loci for PUD and its subtypes (**Supplementary Table 13**). Of those, rs2233580 (*PAX4*; p.R200H; CADD^20^ score = 29.8) was also associated with type 2 diabetes. The variant was common (MAF > 0.05) in 1KG EAS but almost monozygotic in non-EAS populations. rs4745 (p.D159V; PIP = 0.05 for DU; CADD score = 15.2) in *EFNA1* was common in EAS and EUR and was associated with GC. This was the lead variant of cis-splicing quantitative trait loci (sQTL) for *EFNA1* in the stomach and was in high LD with rs4072037^21^ (lead sQTL variant for *MUC1* in the stomach; r^2^ = 0.74 in 1KG EAS). In addition to the missense variants, we found rs4390169 in the credible set (upstream of *EFNA1*; PIP *=* 0.06 for DU; in high LD with rs4745, r^2^ = 0.99 in 1KG EAS and EUR) to be the lead variant of cis-protein QTL (pQTL) in plasma for *EFNA1*^22^.

In the credible sets of previously reported *ABO* and *FUT2* loci for PUD^2,5^, we identified rs8176719 (lead variant at *ABO* locus) and rs1047781 (in the credible sets at *FUT2* locus; PIP = 0.63). rs8176719 deletion resulted in the O allele, whereas rs1047781 (p.I140F) was an EAS-specific common variant (MAF = 0.439 in 1KG EAS), and its A-allele determined the FUT2 secretor status. We performed a logistic regression analysis to investigate the correlation of ABO blood group and FUT2 secretor status with PUD. Blood group O (OR = 1.14, P = 6.0 × 10^−14^) and non-secretor status (OR = 1.17, P = 2.9 × 10^−11^) were significantly correlated with a higher risk of PUD, which was consistent for all PUD subtypes (**Supplementary Table 14**). To investigate the potential interactions between blood group O and non-secretor status, logistic regression analysis including an interaction term was performed. However, significant interactions (P < 0.05/6) were not detected between blood group O and non-secretor status (**Supplementary Table 15**).

### Overlap of eQTL and pQTL with risk variants for PUD

To detect the functionally relevant genes, we searched the GTEx v8 datasets^21^ for overlap of lead cis expression quantitative trait locus (cis-eQTL) with PUD signals or their LD-proxies (LD r^2^ > 0.6 in EAS or EUR)^23^. The most significant eQTL hits for a gene within each tissue type were interpreted (**Supplementary Figure 7**). We identified an overlap of novel variants with eQTLs associated with *IHH, PLCL2, PTGER4, ZNF322, HIATL1, FAM211B*, and *GGT1* in the stomach.

We searched five recent large-scale pQTL datasets^22,24–27^ from serum or plasma for overlap of cis- or trans-pQTL with PUD signals or their LD-proxies. We observed overlaps with 88 unique significant pQTL associations, most of which (93.1%) were trans-pQTL and involved the lead SNP at *ABO* locus (**Supplementary Table 16**). The cis-pQTL alleles in LD with PUD risk alleles were associated with increased levels of EFNA1 and OBP2B, and decreased levels of NHEJ1, ABO, and GGT1 (the cis-pQTLs overlapped with the cis-eQTLs mentioned above for EFNA1, OBP2B, NHEJ1, and GGT1). For trans-pQTLs in LD with the lead variants, we observed links with multiple proteins, including F8, F10, PROS1 (blood coagulation-related), and trefoil factor family peptides (which play important roles in response to gastrointestinal mucosal injury). Additional analysis suggested plausible proteins and pathways (**Supplementary Note**; **Supplementary Table 17**).

### Genetic correlation and pleiotropic effects

We checked cross-trait LD score regression^28^ to evaluate the genetic correlation across PUD-related traits (**Fig. 5a**). DU and GU showed significantly high genetic correlations (rg = 0.79, FDR < 0.05) with each other, as expected. Although not statistically significant, GU showed a positive genetic correlation with GC, whereas DU was negatively correlated. We also investigated the genetic correlation of PUD with dietary habits^29^ and 215 complex traits in BBJ^7^ (**Methods**); no significant genetic correlation was observed between PUD and other complex traits in EAS (FDR < 0.05; **Supplementary Figure 8-10**).

**Fig. 5.**
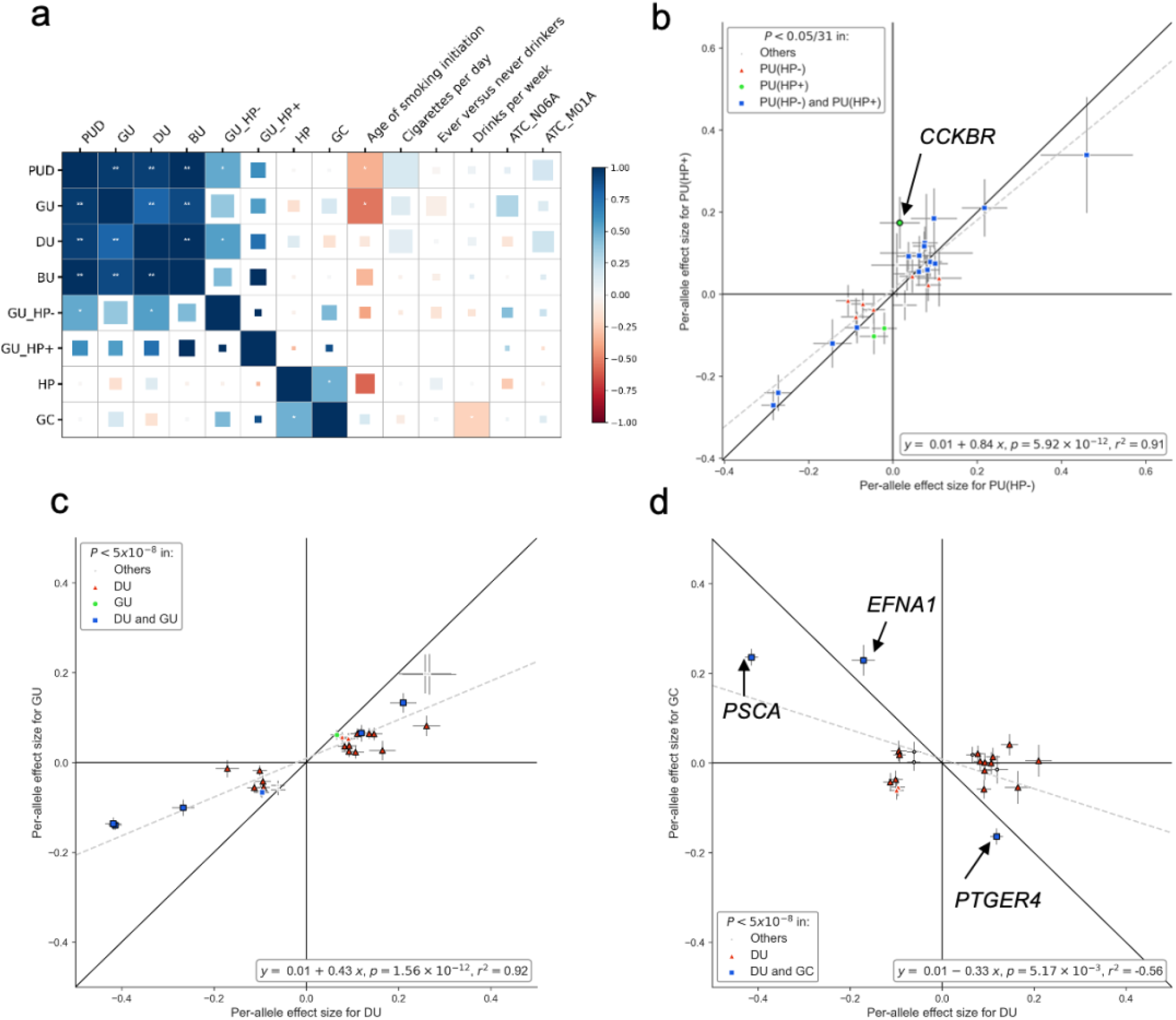
Effect size comparison of distinct variants and genetic correlations across PUD-related traits in the East Asian population. **a**. Genetic correlation among PUD, PUD-related phenotypes, and risk factors. *, P < 0.05; **, FDR < 0.05. The square size in each cell is proportional to –log_10_(P). HP, *H. pylori* infection status determined by anti-*H. pylori* IgG level; GU_HP+, *H. pylori*-positive GU; GU_HP-, *H. pylori*-negative GU. **b**. Effect size comparison for PUD using summary statistics from *H. pylori* stratified analysis. PUD(HP+), *H. pylori*-positive PUD; PUD(HP-), *H. pylori*-negative PUD. **c**. Per-allele effect size (logarithm of odds ratios) comparison using EAS-specific summary statistics for DU and GU. Lead variants and secondary signals associated with PUD or any subtype in the EAS population were selected for comparison. The most significant associations were shown if overlapping variants existed (interval < 500 kb). Only variants with MAF > 0.01 are shown. Black marker edges denote variants with significant heterogeneity (P_het_ < 0.05). The grey dashed line represents the fitted linear regression line. **d**. Effect size comparison between DU and gastric cancer (GC). Effect sizes for DU were obtained from the EAS-specific meta-analysis. GC summary statistics were obtained from previous GWAS conducted in BBJ1.

To investigate the pleiotropic effects of distinct variants, we performed a PheWAS lookup using previous large-scale GWAS in a Japanese population^7^. Among the 27 available lead variants associated with PUD and its subtypes, 16 reached the genome-wide significance threshold for at least one trait (*P* < 5.0 × 10^−8^). From them, 12 variants were associated with at least two traits after Bonferroni correction (*P* < 8.6 × 10^−6^; **Supplementary Figure 11-13**). Both type 2 diabetes (two at *SND1*-*PAX4* locus and one at *GAST* locus) and GC (*EFNA1, PTGER4*, and *PSCA* loci) shared three significant variants after Bonferroni correction (*P* < 8.6 × 10^−6^) with PUD or its subtypes (**Supplementary Figure 14**).

### *H. pylori*-stratified analysis

To examine the differences in genetic architectures between HP-induced and HP-unrelated peptic ulcers, we conducted HP-stratified association tests for PUD in HP-positive and HP-negative individuals from TMM-50K (**Methods**; **Supplementary Table 18**). For the distinct PUD signals identified in the EAS population (29 variants with MAF > 0.01), per-allele effect sizes for PUD between HP-negative and HP-positive status were highly correlated (**Fig. 5b**; slope = 0.84, SE = 0.07, r^2^ = 0.91; **Supplementary Figure 15**). We identified one lead SNP, specifically associated with patients with HP-positive ulcer, at *CCKBR*, and one HP-negative GU locus rs12347577 near *ZNF169* (Cochran’s Q test, P < 0.05). Colocalization analysis corroborated the results. For example, the posterior probability (PP) of only HP-positive PUD having a genetic association at *CCKBR* locus (PP = 0.29) was higher than that for HP-negative PUD (PP = 0.12) (**Supplementary Table 19)**. On the other hand, the most significant loci at *PSCA* were independent of HP infection, marking a strong impact on the onset of PUD regardless of HP infection status.

### Genetic analyses revealed heterogeneity of GU

To further explore the similarities and differences of genetic architecture between GU and DU, we first compared the effect sizes of distinct signals identified in East Asians (lead variants and independent secondary variants) for GU and DU (**Fig. 5c; Supplementary Figure 16**). Notably, the effect sizes for GU showed a strong correlation with those for DU (29 variants with MAF > 0.01; r^2^ = 0.92), which was concordant with the high genetic correlations described above. However, the effect sizes for GU were systematically smaller than those for DU (intercept = 0.01, slope = 0.43, and SE_slope_ = 0.03), with 19 variants (19/29 = 65.5%) showing significant heterogeneity (P_het_ < 0.05) in Cochran’s Q test (**Supplementary Table 20**). To further verify the findings and avoid potential biases for the comparisons, we compared (1) the effect sizes of distinct signals of GWAS in TMM-50K, FinnGen^15^, and UKB^12^, (2) the effect sizes generated by excluding BU samples in the association tests in BBJ1-180K (i.e., no common case in the comparison), and (3) TMM-50K-derived statistics with BBJ-180K-derived statistics (i.e., no common control in the comparison). In any of these comparisons, GU showed high correlation (for variants with MAF > 0.01, r^2^ = 0.75–0.90) with DU, although with smaller effect sizes than DU (**Supplementary Figure 17-19**). Furthermore, we utilized SBasyesS^30^ to estimate the SNP-heritability and polygenicity (defined as the proportion of SNPs with nonzero effects) using Hapmap3^31^ SNPs from EAS-specific summary statistics (**Methods**). Approximately 0.22% of the variants were estimated to have nonzero effects for PUD. Compared to DU, GU showed relatively higher polygenicity but lower heritability (Pi_GU_ = 0.24%, Pi_DU_ = 0.10%; **Supplementary Figure 20**; **Supplementary Table 21**). The results demonstrated that GU and DU showed a high genetic correlation with most risk loci shared and suggested higher heterogeneity of GU^32^.

Finally, using the summary statistics derived from TMM-50K, we generated polygenic risk score (PRS) models with PRS-CS^33^ comprising of 1,029,637 variants and tested the PRS in BBJ1-180K for associations with PUD or PUD subtypes to investigate the genetic overlap among PUD and PUD subtypes. Compared to HP-positive PRS, HP-negative PRS generally showed stronger associations with PUD or PUD subtypes. The strongest association was between DU PRS and DU (OR = 1.229, confidence interval, CI = 1.20–1.25). DU PRS showed a stronger association with GU than GU PRS (OR = 1.04, CI = 1.03–1.06). **(Supplementary Table 22)**. The results further validated that GU shares risk loci with DU while having higher heterogeneity than the latter^34^.

### Comparison of the effect estimates between PUD subtypes and GC

Considering that DU is a protective factor against GC, we compared the effect sizes of distinct signals in EAS between PUD subtypes and GC to investigate the genetic factors potentially contributing to the observation. Summary statistics for GC in EAS were obtained from a previous study in BBJ1-180K, which included approximately 6,500 cases^34^ (**Methods**). For 23 available variants existing in both datasets, we found the effect sizes for DU to be negatively correlated with that for GC (slope = -0.33, SE_slope_ = 0.10; **Fig. 5d; Supplementary Table 23; Supplementary Figure 21**). It was noteworthy that lead variants linked to *EFNA1* (Ephrin A1, a member of the EFN family), *PTGER4* (receptor for prostaglandin E2, PGE_2_), and *PSCA* showed relatively strong but opposite effects on DU and GC **(Supplementary Table 23**), which suggested that the alleles of these variants, which increased the risk for PUD, could decrease the risk for GC. When removing the three variants from the regression, negative correlation was not observed for the 20 variants (slope = -0.06, SE = 0.10; **Supplementary Figure 21**), indicating that negative correlation between DU and GC was largely affected by the three variants.

### Gene-based and gene-set analysis

Gene-level analysis using MAGMA^35,36^ (**Methods**) detected 29 genes significantly associated with PUD (*P* < 6.5 × 10^−7^; **Supplementary Table 24**), 45 genes associated with DU, and 15 genes associated with GU, in the EAS population. In total, 47 distinct genes were associated with PUD or PUD subtypes. Multiple genes identified in gene-level analysis were reportedly related to GC (*PTGER4*^37^, *PRKAA1*^38^, *GNAQ*^39^, *GNAS*^40^, *NHEJ1*^41^, *IHH*^42^, and *JUP*^43^). Based on the gene-level statistics, we additionally performed pathway enrichment analysis and identified one gene set after Bonferroni correction (nikolsky_breast_cancer_8q23_q24_amplicon, including genes within amplicon 8q23-q24 identified in a study of breast tumors^44^) (*P* < 8.0 × 10^−7^; **Supplementary Table 25**).

### Tissue- and cell-type specificity analysis

We tested the tissue-level specificity employing MAGMA^36^ with GTEx v8 datasets^21^ in EAS individuals to investigate the tissue types related to PUD and its subtypes. Significant genetic enrichments (FDR < 5%) were observed in the stomach, pancreas, small intestine, and kidney for PUD, in the stomach, pancreas, and prostate for DU, and in the stomach for GU (**Fig. 6a**).

**Fig. 6.**
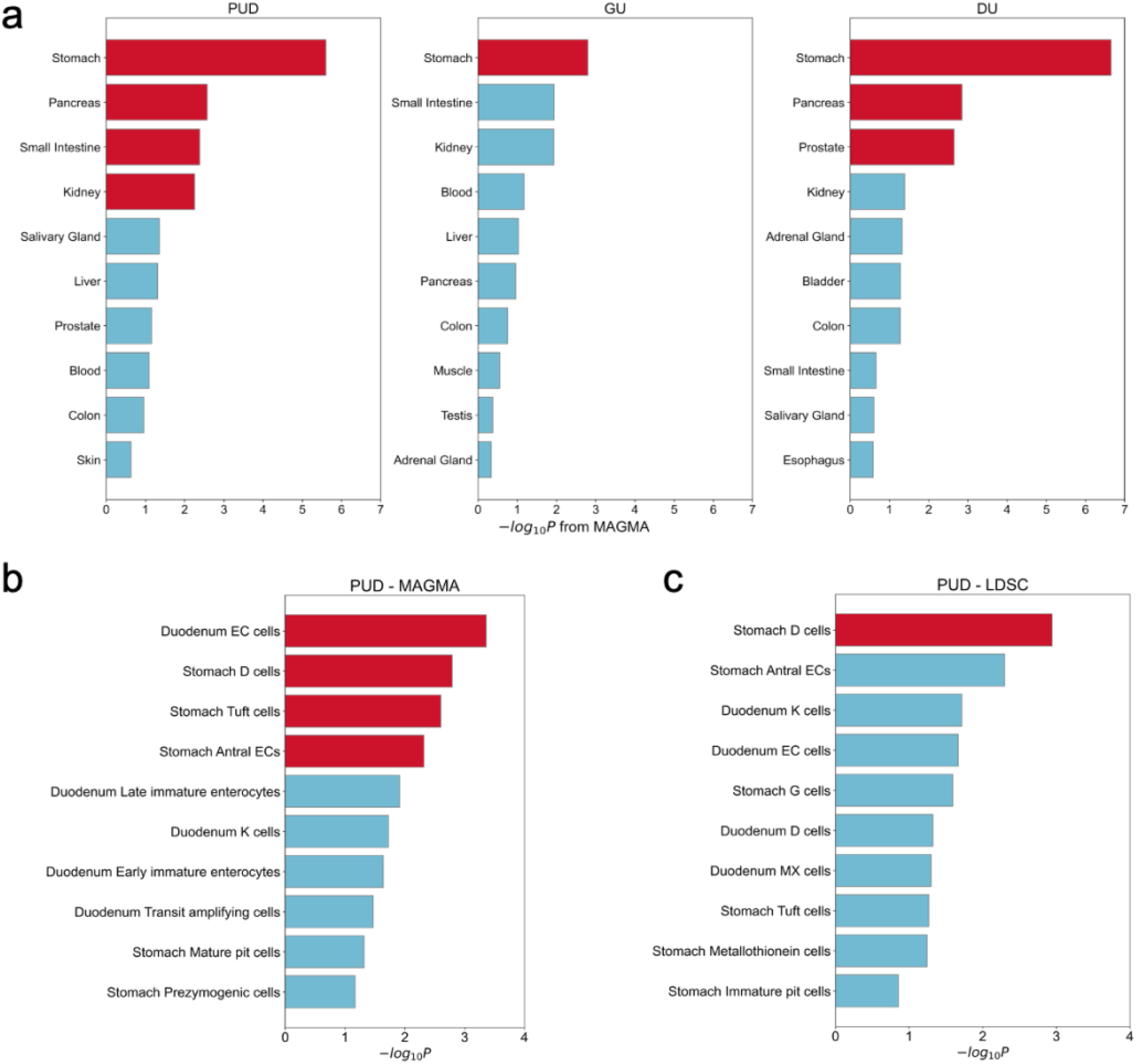
Tissue- and cell-type specificity analysis. **a**. Associations across peptic ulcer phenotypes and 30 general tissue types were analyzed using MAGMA with East Asian-specific summary statistics and the GTEx version 8 dataset. **b, c**. Associations between PUD and cell types in the stomach and duodenum were analyzed using MAGMA and LDSC (testing for enrichment of the 10% most specific genes in each cell type). Inverse variance weighted meta-analysis combined statistics from East Asian and European ancestries for each method. x-axis,–log_10_(P) derived from meta-analyzed estimates. **a, b, c**. Red bars imply significant associations after corrections (FDR < 5%). Only the ten most significant associations for each phenotype are shown.

To further characterize specific cell types associated with PUD in the gastric and duodenal tissues, we utilized publicly available single-cell RNA sequencing (scRNA-seq) datasets of the human stomach and duodenum^45^. Top 10% of the most specific genes for each cell type were selected as the cell-specific gene sets (**Methods**). We performed cell specificity analysis using LDSC^46^ and MAGMA in EAS and EUR individuals, respectively. To increase statistical power, we conducted a fixed-effect meta-analysis combining EAS and EUR results for each method (**Methods**). For PUD, we found that stomach D cells reached the significance threshold (FDR < 5%) in both the analyses of MAGMA and LDSC (**Fig. 6b-c**). Additionally, duodenal enterochromaffin cells (EC cells), stomach antral ECs, and stomach tuft cells were significantly (FDR < 5%) associated with PUD, as per MAGMA (**Fig. 6b**). Somatostatin produced by stomach D cells inhibits the secretion of a variety of gastrointestinal hormones, including the gastrin secreted by stomach G cells that stimulate gastric acid secretion. EC cells secrete serotonin (5-HT, a neurotransmitter) with diverse gastrointestinal functions, and tuft cells (chemosensory epithelial cells) secrete interleukin-25 (IL-25) driving type 2 immune response to parasitic infection. Together, the findings suggested the important role of gastrointestinal hormone regulation and immune response in PUD etiology (**Supplementary Table 26-27; Supplementary Figure 22-27)**.

## Discussion

Our GWAS meta-analyses of PUD and PUD subtypes discovered 33 autosomal susceptibility loci, of which 25 had not been reported in previous GWAS (19 in East Asian-specific analysis and 6 in cross-ancestry analysis). The loci were mostly shared across ancestries with strong correlations of effect sizes. Our cross-ancestry analysis emphasized the high genetic correlation of DU and suggested the heterogeneity of GU across ancestries. The larger effect sizes of *MUC1* and *MUC6* in populations of EUR ancestry are in reasonable agreement with their critical roles in the protection from NSAID-induced injury, given the much lower prevalence of HP infection in western countries compared to that in East Asian populations^47^.

Multiple novel loci (*PAX4*^48^, *PDX1*^49^, *IHH*^50^, and *SLC22A3*^51^) and reported loci (*CCKBR, CDX2*, and *GAST*) were found to be related to cell differentiation and gastrin signaling. By integrating scRNA-seq datasets, we identified the association of PUD with certain hormone-secreting cells, including stomach D cells (somatostatin), stomach antral and duodenal EC cells (serotonin), and stomach tuft cells. HP infection increases gastrin secretion^52^; our results showed the signal at *CCKBR* (receptor for gastrin) to be HP-dependent. PUD risk allele of the lead SNP (rs12792379) is in LD with the eQTL allele associated with higher *CCKBR* expression in multiple tissues^21^, including esophagus mucosa. Taken together, our results provided genetic evidence of gastrointestinal cell differentiation and hormone regulation being critical in PUD etiology.

As expected, we observed high genetic correlation between GU and DU; effect size comparisons demonstrated that GU shared risk loci with DU, but had smaller effect sizes than DU. Polygenicity of GU was higher than that of DU. SNP-based heritability estimate for DU (liability-scaled) was almost twice as high as for GU. Additionally, DU PRS showed a stronger association with GU than GU PRS in East Asians. The results revealed the genetic difference between GU and DU and reflected a higher heterogeneity of GU^53–55^.

We found three variants (linked to *EFNA1, PTGER4*, and *PSCA*) to potentially contribute to the protective role of DU against GC. *EFNA1* suppresses tumor growth while PGE_2_ supports tumor growth by promoting angiogenesis^37,56^. PUD risk alleles resulted in increased levels of *EFNA1* and reduced levels of *PTGER4*, whereas GC risk alleles were associated with a decreased level of *EFNA1* and increased level of *PTGER4*. This suggested that the risk alleles of *EFNA1* and *PTGER4* for GC (non-risk alleles for PUD) potentially benefited peptic ulcer healing while imposing an increased risk for GC through up-regulated cell proliferation and angiogenesis. Additionally, we also detected multiple PUD-risk cancer-related genes (e.g., *IHH, GNAS, NHEJ1, JUP*, and *MECOM*), which provided potential targets contributing to the different outcomes of PUD or GC.

Although we identified multiple associations, the current study has several potential limitations. First, the phenotypic information of PUD and subtypes was obtained via interviews and reviews of medical records. However, the prevalence rate of PUD was consistent with that in previous epidemiological studies, and our study replicated most of the previously identified loci. Second, due to the lack of information regarding the chronological order of PUD onset and anti-inflammatory drug use at PUD onset in this study, the specific interaction of NSAIDs with host genetic factors was under-explored. Third, detailed information on the anatomic site of the ulcers or the strains of HP was not available. Even though our subtype analysis revealed the overall similarities and differences in genetic architecture, large sample size and more detailed classifications are still warranted to elucidate the potential heterogeneity further.

In summary, the current study tripled the number of risk loci for PUD and its subtypes, and improved our understanding of the genetic architecture of PUD. The findings provided insight into the biological pathways involved in PUD pathogenesis and potential links between PUD and GC. We demonstrated that besides *H. pylori*-related loci, host genetic factors potentially involved in gastric hormone regulation, cell differentiation, and proliferation might play important roles in PUD pathogenesis. Our single cell analysis further revealed the association of serotonin-secreting EC cells, somatostatin-secreting stomach D cells, and stomach tuft cells with PUD, indicating their key role in PUD etiology.

## Data Availability

Summary statistics, including the results of GWAS at the discovery stage and that of the East Asian-specific and cross-ancestry meta-analysis in this study, will be available at National Bioscience Database Center (NBDC, https://humandbs.biosciencedbc.jp/) Human Database or Japanese ENcyclopedia of GEnetic associations by Riken (JENGER, http://jenger.riken.jp/) website upon its acceptance by a journal.
Genotype data for BBJ were deposited at NBDC Human Database (BBJ1-180K, research ID:hum0014; BBJ1-12K and BBJ2-42K, research ID:hum0311).

## Online methods

### Study participants

BioBank Japan Project (BBJ, https://biobankjp.org/en/)^10^, a hospital-based study, was founded in 2003, and enrolled approximately 200,000 participants of mainly Japanese ancestry, with at least one of 47 common diseases from 2003 to 2007, as the first cohort (BBJ1). From 2013 to 2017, BBJ additionally collected DNA and clinical information from 67,321 newly registered participants with at least one of 38 common diseases as the second cohort (BBJ2).

Genotype data of the case and control individuals included in the discovery-stage GWAS were obtained from the primary dataset of BBJ1, including 181,927 individuals (denoted as BBJ1-180K). Clinical information, including age and sex, was obtained from clinical records of BBJ-participating hospitals. In this study, we included samples of age ≥ 18 years. We performed principal component analysis (PCA) using 1000 Genome^11^ samples and then projected all samples onto the same space. We excluded outliers from the East Asian cluster.

Replication was conducted in three Japanese studies, namely an additional and independent set of BBJ1 cohort of 11,715 individuals (denoted as BBJ1-12K in this study, which was not included in the BBJ1-180K), a cohort of 42,689 individuals from BBJ2 (denoted as BBJ2-42K), and a population-based cohort of 49,621 individuals from Tohoku University Tohoku Medical Megabank (TMM; https://www.megabank.tohoku.ac.jp/english/) Project (denoted as TMM-50K)^13^.

The clinical characteristics of these cohorts are provided in detail in **Supplementary Table 1**. The research project was approved by the ethics committees at the Institute of Medical Science, the University of Tokyo (application number 29-74-A0215), and Iwate Tohoku Medical Megabank Organization, Iwate Medical University (application number HG H25-2).

### Phenotype definition

In this study, we assessed peptic ulcer disease (PUD), which is a combination of the two major subtypes, namely duodenal ulcers (DU) and gastric ulcers (GU). PUD cases were obtained from the combination of individuals with any of the two major PUD subtypes (DU and GU). Clinical information for GU and DU cases was obtained via interviews and reviews of medical records using a standardized questionnaire in BBJ1-180K, BBJ1-12K, and BBJ2-42K, and by self-administered questionnaires in TMM-50K^57^.

Individuals with comorbidities of GU and DU were additionally categorized into a group BU. Individuals without a given diagnosis of peptic ulcers or any *H. pylori*-related diseases (GU, DU, or gastric cancer) were used as control samples (**Supplementary Table 1**).

### Genotyping and imputation

All samples included in the discovery-stage GWAS (i.e., BBJ1-180K) were genotyped with either Illumina HumanOmniExpressExome BeadChips or a combination of Illumina HumanOmniExpress and HumanExome BeadChips. Samples from the BBJ1-12K and BBJ2-42K cohorts were genotyped with Infinium Asian Screening Array BeadChips. Samples from the TMM-50K cohort were genotyped with Axiom Japonica Array JPAv2 (Thermo Fisher Scientific, MA, USA). In the BBJ1-180K dataset, samples with call rates < 98% were excluded. For QC of autosomal genotypes, we excluded the genotyped variants based on the following criteria for BBJ1-180K: (1) call rate < 99%, (2) heterozygote count < 5, (3) Hardy–Weinberg-equilibrium P < 1.0 × 10 ^*−6*^, and (4) concordance rate < 99.5% or non-reference discordance rate ≥ 0.5% between array genotypes and whole-genome-sequence dataset using overlapping participants (n = 939), as described previously^58^. We applied the same criteria (except (4)) as in the discovery stage to the replication sets of BBJ1-12K and BBJ2-42K. Additionally, we excluded the samples with amyotrophic lateral sclerosis in BBJ1-12K due to its comparatively high proportion. In the replication using TMM-50K, we excluded samples with call rate < 99% and variants with (1) call rate < 99%, (2) heterozygote count < 5 or (3) Hardy–Weinberg-equilibrium P < 1.0 × 10 ^-6^. For QC of variants on chromosome X, we excluded genotyped variants with (1) call rate < 99% in males, females, or both, and (2) Hardy–Weinberg-equilibrium P < 1.0 × 10 ^*−*6^ in females.

Pre-phasing was conducted using Eagle2 (v2.4.1; https://alkesgroup.broadinstitute.org/Eagle/)^59^. Imputation was performed with Minimac4 (v1.0.2; https://github.com/statgen/Minimac4) using the 1000 Genomes Project Phase 3^11^ version 5 (1KGp3v5) ALL panel (https://genome.sph.umich.edu/wiki/Minimac3#Reference_Panels_for_Download). For chromosome X, haplotypes were pre-phased for males and females, and variants were imputed separately in males and females using the same software. Imputed variants with Rsq < 0.3 were excluded in the association analysis. More than 13 million variants were included in the discovery-stage association analysis.

### Genome-wide association analysis

Single-variant association analysis was performed with SAIGE (v0.44; https://github.com/weizhouUMICH/SAIGE)^12^, which implements a generalized mixed model with SPA correction controlling for case-control imbalance and cryptic relatedness. The regression included age, sex, and top 10 PCs as covariates. For step 1, LD-pruned genotyped variants (PLINK --indep-pairwise 50 5 0.2, https://www.cog-genomics.org/plink/)^60^ with MAF > 1% were used to estimate the null models with leave one chromosome out (LOCO). Variants with MAC < 20 were excluded from the association tests (step 2). For sex-stratified analysis, single-variant association analyses were performed with SAIGE adjusting for the same set of covariates other than sex in males and females.

For association tests of the X chromosome, variants were tested separately in males and females using the corresponding null models estimated by autosomes in each sex. Haploid-based dosages of the non-pseudo autosomal (non-PAR) region of males were multiplied by 2. The results for each sex were then meta-analyzed using inverse-variance weighted methods implemented in METAL software (v2011-03-25, http://csg.sph.umich.edu/abecasis/Metal/index.html)^61^.

Genome-wide significant loci were determined by iteratively extending 500-kb flanking regions around the most significant variant until no genome-wide significant variant (P < 5.0 × 10 ^*−*8^) was detected within the extended regions. The most significant variant in each locus was selected as the lead variant. Loci for different traits with lead variants within 500 kb of each other were considered the same, denoted by the most significant lead variant from the locus. Significant variants in the MHC region (GRCh37, chromosome 6: 25–34 Mb) were counted as one locus due to complexity of the region.

### LD score regression

We performed LD score regression (v1.0.0; https://github.com/bulik/ldsc)^14^ to examine the bias caused by confounding factors, such as population stratification or cryptic relatedness. We employed the LD scores provided by authors for the East Asian population, which were estimated from 1000 Genomes Project EAS individuals. To convert observed-scale heritability to liability-scale heritability, the population prevalence rates in East Asian populations were set to 6.2%, 6.9%, 10.8%, and 1.8% for DU, GU, PU, and BU, respectively. The prevalence rates were estimated from the population-based TMM-50K and were similar to those in previous epidemiological studies^1^.

### Replication of significant associations and EAS-specific meta-analysis

We compared the directions and effect sizes with the replication GWAS sets for the lead variants of significant loci identified in the discovery-stage GWAS. The results of GWAS at the discovery and replication stages were combined using the fixed-effect inverse-variance method implemented in METAL. Heterogeneity was estimated by Cochran’s Q test. We considered the lead variants identified in discovery-stage GWAS as replicated if the variants reached nominal significance threshold (P_rep_ < 0.05) in the same direction in at least two of the replication GWAS.

### Cross-ancestry meta-analysis

Summary statistics of PUD and PUD subtypes for European individuals were obtained from FinnGen (release 6 for PUD, DU, and GU; https://www.finngen.fi/en/access_results)^15^, a published GWAS of PUD in UKB (https://cnsgenomics.com/content/data)^5^ and PheWeb UKB-SAIGE^12^ (DU and GU; https://pheweb.org/UKB-SAIGE/) (Details in Supplementary Table 7).

Genome coordinates of summary statistics were converted from GRCh38 (hg38) to GRCh37 (hg19) using UCSC LiftOver tool^62^ if the original summary statistics were based on GRCh38. We performed additional quality control and harmonization for all summary statistics prior to meta-analyses; only autosomal variants in the 1KGp3v5 dataset were included in the meta-analysis; all variants were normalized^63^, duplicate and multiallelic variants were removed for each dataset, and variants with imputation quality scores less than 0.3 were removed. For summary statistics from UKB-SAIGE (imputation quality scores not available), variants included in the previously published GWAS of PUD in UKB were kept, missing information of chromosome and base pair positions were assigned according to rsID, variants with extreme effect size values (log(OR) < −10 or log(OR) > 10) were removed, variants with minor allele count (MAC) < 5 were removed, and the strand of palindromic variants with MAF < 0.40 was further inferred using the allele frequencies obtained from each population in 1KGp3v5 dataset. Finally, we compared the effect allele frequencies in summary statistics and the population-specific alternative allele frequencies in 1KGp3v5. Variants with deviation in allele frequencies > 0.16 were excluded. In total, more than 19 million variants were included in the meta-analysis.

The fixed-effect inverse-variance method was used to conduct meta-analyses integrating GWAS results in EAS and EUR populations using METAL. We additionally performed fixed-effect meta-analyses to generate EUR-specific summary statistics using the two EUR datasets. Population-specific meta-analyses were used to compare the effect sizes of lead variants identified in cross-ancestry meta-analyses between EAS and EUR populations.

MR-MEGA (v0.2;http://www.geenivaramu.ee/en/tools/mr-mega)^16^ was used to perform cross-ancestry meta-regression with four axes of genetic variation derived via multi-dimensional scaling. P values were recalculated using chi-squared statistic due to the lack of support in MR-MEGA for P < 1.0 × 10 ^*−*14^.

### Genetic correlation estimation

To assess the genetic correlation between PUD and common binary traits and quantitative traits in East Asian populations, we used cross-trait LDSC^28^ with LD scores estimated from 1KG EAS individuals. East Asian summary statistics were obtained from previous GWAS in BioBank Japan^29,34^. MHC region was excluded.

To evaluate the cross-ancestry correlations of genetic effect for PUD and subtypes between EAS and EUR, Popcorn (v1.0; https://github.com/brielin/Popcorn)^17^ was used with pre-computed cross-ancestry LD scores estimated from 1000 Genomes Project EUR and EAS populations. For these analyses, meta-analyzed summary statistics of PUD in EAS and EUR for HapMap 3 SNPs (without MHC region) were used.

### Blood group and secretor status interaction analysis

ABO blood groups for unrelated individuals used in discovery-stage GWAS in BBJ1-180K (KING kingship coefficient^64^ < 0.0884, N = 164,613) were inferred from two genotyped variants described previously ^2^. Secretor status was inferred using the best-guess genotype of imputed variants rs1047781 (p.Ile140Phe)^65^, where AA or AT genotypes are secretors, and TT are non-secretors. Logistic regression analyses adjusting for age, sex, and top 10 PCs were performed to examine the association of blood group or secretor status with PUD or subtypes. Blood group-specific effect sizes were estimated using the target blood group as exposure and combination of the other three blood groups as non-exposure (for example, individuals with A vs. B, AB, and O). Secretor status effect sizes were estimated considering the non-secretor status as exposure and the secretor status as non-exposure. To investigate the interaction of blood group O with non-secretor status, we further performed logistic regression analyses for blood group O–secretor interaction, adjusting for O blood group, FUT2 secretor status, age, sex, and top 10 PCs. Similarly, the blood group O – non-secretor status interaction was tested using imputed dosages of the variants determining O antigen and secretor status. All the above-mentioned logistic regressions were conducted in R v4.1.0. Additionally, we explored a previous pQTL study that had investigated proteomic associations with ABO blood groups and FUT2 secretor status for proteins associated with secretor status and A, B, and AB blood groups.

### Conditional analysis by COJO

GCTA-COJO (v1.93.2; https://yanglab.westlake.edu.cn/software/gcta/#COJO)^18^ was employed to perform conditional analysis in each significant locus identified in EAS-specific meta-analysis of PUD and subtypes. We constructed an LD reference panel using the best-guess imputed genotype of 20,000 randomly selected and unrelated individuals of East Asian ancestry from BBJ1-180K. Stepwise model selection was conducted first to select independent association signals (P < 5.0 × 10^*−*8^), and a joint analysis of these selected signals was performed next. Variants with MAF > 0.01 were included in the analysis.

### Fine-mapping and variant annotation

Fine-mapping was conducted using SuSiE (v0.11.92; https://github.com/stephenslab/susieR/)^19^ with default configurations while allowing ten putative causal variants within each locus. Unrelated individuals (KING kingship coefficient value < 0.0884, N = 171,085) from BBJ were used as LD reference, computed by LDstore (v2.0; http://www.christianbenner.com/)^66^ based on the imputed dosages.

We defined regions based on the 3 Mb window centered at the lead variants and merged them if the window overlapped. Only variants with Rsq ≥ 0.5 were included in fine-mapping. We reported the credible sets (CS), which have a 95% probability of harboring one causal variant.

Variants identified in the GWAS were annotated using ANNOVAR (v2020-06-07; -protocol refGene,avsnp150,clinvar_20200316; https://annovar.openbioinformatics.org/en/latest/)^67^.

### *H. pylori*-stratified analysis

To investigate the interaction of HP with host genetic factors for the development of peptic ulcers, we performed HP-stratified analyses in HP-positive and HP-negative individuals from TMM-50K. *H. pylori* infection status was determined by anti-HP serum IgG antibody, measured by the latex agglutination immunoassay (LIA). Individuals with anti-HP serum IgG antibody titer ≥ 10 U/mL were categorized as HP-positive. Association tests were performed with the same settings as in the discovery-stage GWAS. Cochrane Q and I^2^ statistics (calculated by R package metafor v3.4, https://www.metafor-project.org/doku.php)^68^ were used to test the effect size heterogeneity between *H. pylori* -positive and *H. pylori*-negative GWAS for each subtype.

Colocalization analysis was conducted using coloc package (v5.1.0; https://chr1swallace.github.io/coloc/)^69^ for each significant locus identified in EAS meta-analysis under a single causal variant assumption.

### Estimation of polygenicity using SBayesS

To estimate the polygenicity (defined as the proportion of SNPs with nonzero effects) and the strength of negative selection (defined as the relationship between MAF and effect sizes, and denoted by S) for PUD, we utilized SBayesS from GCTB software (v2.0; https://cnsgenomics.com/software/gctb/#Overview)^30^. SBayesS employs a Bayesian mixed linear model and reports SNP-based heritability, polygenicity estimates, and a metric that indicates negative selection.

An LD reference panel for EAS was constructed using the approach described previously ^30^. Briefly, a full LD matrix on HapMap3 SNPs was computed using 50,000 randomly selected and unrelated East Asian individuals from BBJ1-180K. The off-diagonal entries of full LD matrix were shrunk with the interpolated genetic map for the 1000 Genomes Project JPT population (https://github.com/joepickrell/1000-genomes-genetic-maps). The effective population size and genetic map sample size were set to 11,600 and 100, respectively, according to 1000 Genomes Project phased OMNI data (http://ftp.1000genomes.ebi.ac.uk/vol1/ftp/technical/working/20130507_omni_recombination_rates/). The sparse shrunk LD matrix used for SBayesS was created by setting elements of the shrunk matrix to zero if their chi-squared statistic under the sampling distribution of the correlation coefficient did not exceed 10.

For a cross-trait comparison of polygenicity estimates in EAS, we included 42 binary traits of BBJ1 in the analysis^34^.

We ran four parallel MCMC chains with a length of 50,000 and a burn-in size of 20,000 for each trait. To evaluate the convergence in MCMC, potential scale reduction statistics for each parameter was computed. Traits with potential scale reduction statistic < 1.2 for all three parameters, including SNP-based heritability, polygenicity estimates, and S, were considered to have good convergence, and were, therefore, used in the study.

### Polygenic risk score construction and evaluation

Polygenic risk score (PRS) models for PUD and PUD subtypes were constructed in East Asians. We used the summary statistics derived from replication GWAS and HP-stratified analysis in the population-based Japanese cohort TMM-50K. PRScs (v2021-Jun-4, https://github.com/getian107/PRScs)^33^ were employed to compute PRS models using HapMap3 SNPs with an EAS-specific LD reference panel from the 1000 Genomes Project. Global shrinkage parameters were obtained from the data by PRScs using a fully Bayesian approach (PRS-CS-auto). We applied the models in BBJ-180K and then tested the associations of PRS with PUD and PUD subtypes using logistic regression adjusted for age, sex, and top five PCs. We evaluated the predictive ability of each PRS model by its improvement of AUC over a base model that includes age, sex, and top five PCs.

### Gene-based analysis and pathway analyses

Gene-based and pathway analyses were performed using MAGMA (v1.08; https://ctg.cncr.nl/software/magma)^35^ implemented in FUMA (v1.3.8; https://fuma.ctglab.nl/)^36^. An LD reference panel constructed from 1000 Genomes EAS population was used. A total of 19,033 protein-coding genes (ENSEMBL^70^ v92) were tested. Results of the gene-based analysis were then employed to conduct gene-set enrichment analysis with a total of 15,485 curated gene sets, and GO terms from MsigDB^65^ v7.0 were tested for association.

### Per-allele effect size comparison

For pair-wise effect size (logarithm of odds ratios) comparison among PUD, PUD subtypes, and gastric cancers in the EAS population, we selected the non-overlapping (interval between adjacent variants > 500 kb) lead variants identified in EAS-specific meta-analysis and the independent signals identified in COJO analysis. For loci associated with two or more phenotypes, we selected the most significant associations (lead variants with the lowest P-value) for comparison. Effect sizes in EAS-specific meta-analysis were used for PUD and PUD subtypes. Summary statistics for GAC were obtained from a previous study in BBJ1-180K^13^. For cross-ancestry comparison of variant effect sizes, we included all non-overlapping lead variants associated with PUD or any subtypes in population-specific meta-analysis or cross-ancestry meta-analysis. Associations, with the lowest P values, of loci associated with more than one phenotype, were selected for comparison and effect sizes in the population-specific meta-analysis were used. Cochrane Q test was used to test heterogeneity across the effect sizes.

### PheWAS in BBJ

To investigate whether variants associated with PUD were also associated with other human complex traits in EAS, statistics of the non-overlapping lead variants and secondary signals for 215 case-control and quantitative traits were obtained from BioBank Japan PheWeb (https://pheweb.jp/)^7^. LD proxy (r^2^ > 0.6) with the highest r^2^ estimated from 1KG EAS was used if a variant was unavailable in the datasets. After multiple-test correction, the significance threshold was set to P < 8.6 × 10^*−*6^ (0.05/215/27).

### Tissue/cell-type specificity analysis

MAGMA^35^ gene-property analysis implemented in the SNP2GENE method of FUMA^36^ was employed for tissue-type specificity analysis with gene expression profile from GTEx v8 dataset^21^. A total of 54 non-diseased tissue types and 30 general tissue types were tested. Tissue types with a false discovery rate (FDR) < 5% were considered significant.

To identify the cell types associated with PUD in the stomach and duodenum, processed single-cell RNA sequencing datasets of the human stomach and duodenum were obtained from a previous study^45^, which filtered for cells with more than 1,500 transcripts per cell and genes expressed by at least three transcripts in at least one cell. A total of 13,980 genes for 19 cell types in the stomach and 17 cell types in the duodenum were included in the study. The top 10% most specifically expressed genes based on fold-change (defined as the average transcript counts of all cell types except the target cell type divided by the average transcript counts of the target cell type) were extracted for each cell type. SNPs in cell-type-specific genes were used to compute partitioned LD scores in 1KG phase 3 EAS or EUR population. The gene coordinates were extended by a window size of 100 kb to capture the effects of regulatory elements. Stratified LD score regressions^46^ were performed using the partitioned LD scores of cell-type-specific genes, partitioned LD scores of all available genes in the dataset, and the baseline model of 53 annotations for each ancestry on HapMap3 SNPs excluding the MHC region (downloaded from https://alkesgroup.broadinstitute.org/LDSCORE/).

We performed gene-set enrichment analysis using MAGMA with the cell-type-specific gene sets described above. We used 1000 Genomes Project phase 3 EAS and EUR population datasets as reference panels. Variants with MAF < 0.01 and MHC regions were excluded from the analysis. The gene coordinates were extended by window sizes of 35 kb upstream and 10 kb downstream to capture the effects of regulatory elements. IVW meta-analysis was performed using statistics of both ancestries for each method to increase statistical power. P values were calculated using the one-tailed test. Cell types with a false discovery rate (FDR) < 5% within each expression dataset were considered significant.

### eQTL and pQTL analysis

To characterize the effect of variants on gene expression level, we extracted LD proxies (with r^2^ > 0.6) in EAS or EUR 1KG Phase 3 with the lead variants and secondary signals in EAS. We extracted only significant SNP-gene pairs with FDR < 0.05 (pre-computed by the authors) from GTEx version 8^21^. We checked the overlap between the lead variants and secondary signals (including proxies) and cis-eQTL variants in GTEx v8. The most significant cis-eQTL association for each gene in each tissue was selected for interpretation.

To characterize the effect of variants on protein level, we extracted LD proxies (with r^2^ > 0.6) in EAS or EUR 1KG Phase 3 with the lead variants and secondary signals in EAS. We extracted genome-wide significant SNP-protein associations from five published large-scale pQTL studies in recent years^22,24–27^, conducted in individuals of mainly European ancestry. We then checked the overlap between the lead variants (including LD proxies) with cis- and trans-pQTL. The most significant association for each protein was selected for interpretation.

## Data availability

Summary statistics, including the results of GWAS at the discovery stage and that of the East Asian-specific and cross-ancestry meta-analysis in this study, will be available at National Bioscience Database Center (NBDC, https://humandbs.biosciencedbc.jp/) Human Database or Japanese ENcyclopedia of GEnetic associations by Riken (JENGER, http://jenger.riken.jp/) website upon its acceptance by a journal. Genotype data for BBJ were deposited at NBDC Human Database (BBJ1-180K, research ID:hum0014; BBJ1-12K and BBJ2-42K, research ID:hum0311)

## Acknowledgments

We want to acknowledge all the participants and investigators of BioBank Japan, Tohoku Medical Megabank, UK Biobank, and FinnGen. We express our gratitude to Prof. Mark Lathrop for his valuable support. This research was supported by the Ministry of Education, Culture, Sports, Sciences and Technology (MEXT) of Japanese government and the Japan Agency for Medical Research and Development (AMED) under grant numbers JP21km0605001 (the BioBank Japan project), JP15km0105004 and JP21tm0124006 (the Tohoku Medical Megabank Project, Special Account for the Reconstruction of the Great East Japan Earthquake), JP20km0405001 (for the supercomputer resource), and JP21km0405215 (to C.T., K.M., and Y.K.) We thank the Human Genome Center, the Institute of Medical Science, the University of Tokyo for providing the super-computing resource used in this study.

