## Supplementary Materials for "East Asian-specific and cross-ancestry genome-wide meta-analyses provide mechanistic insights into peptic ulcer disease"

### Supplementary Notes

#### Plausible pathways suggested by GWAS and pQTL analysis.

By searching for pQTL associations[1–5], we found that the PUD risk alleles of lead variants at *ABO* and *GGT1* were linked with a reduced level of coagulation factor VIII (F8) and increased levels of factor X (F10) and PROS1(cofactor to activated protein C in the degradation of factor VIII). The risk allele of a lead SNP (rs1801020; F12) identified in the cross-ancestry meta-analysis is associated with decreased plasma levels of factor XII[6]. Factor XII, VIII, X, and PROS1 are all involved in the intrinsic pathway of blood coagulation[7,8]. These suggest that blood coagulation may be involved in peptic ulcer bleeding and healing. It is also likely that these signals were identified due to selection bias since PUD patients with severe symptoms are more likely to be detected than those without such severity. Additionally, in the pQTL study[1] that observed the proteomic associations with ABO blood groups and FUT2 secretor status, we found 31 proteins to be significantly associated with secretor status and non-O blood groups in the same direction (**Methods**). Notably, five proteins (CBLIF, CRNN, DSG2, REG1A, and REG1B) showed directionally concordant associations with secretor status and all three non-O blood groups (A, B, and AB) (**Supplementary Table 17**).

### Supplementary Figures

**Table of contents**

Supplementary Figure 1. Study workflow. 3
Supplementary Figure 2. Venn diagram of cases used in the discovery-stage GWAS. 4
Supplementary Figure 3. Manhattan plot and Q-Q plot for PUD and PUD subtypes from the discovery stage GWAS in BBJ1-180K. 4
Supplementary Figure 4. Manhattan plot and Q-Q plot for PUD and PUD subtypes from the East Asian-specific meta-analysis. 5
Supplementary Figure 5. Manhattan plot and Q-Q plot for PUD and PUD subtypes from the cross-ancestry meta-analysis. 6
Supplementary Figure 6. Cross-ancestry effect size comparison of lead variants for PUD and PUD subtypes. 7
Supplementary Figure 7. Overlap between PUD signals and significant cis-eQTL variants of the GTEx database. 8
Supplementary Figure 8. Genetic correlation heatmap of PUD and dietary habits in East Asian population. 9
Supplementary Figure 9. Genetic correlation heatmap of PUD and quantitative traits in East Asian population. 9
Supplementary Figure 10. Genetic correlation heatmap of PUD and binary traits in East Asian population. 9
Supplementary Figure 11. PheWAS heatmap of PUD risk variants with ATC codes and quantitative traits. 10
Supplementary Figure 12. PheWAS heatmap of PUD risk variants with binary traits (Part 1/2). 11
Supplementary Figure 13. PheWAS heatmap of PUD risk variants with binary traits (Part 2/2). 12
Supplementary Figure 14. Summary of significant associations identified in PheWAS. 13
Supplementary Figure 15. Effect size comparison of lead variants and secondary signals for H.pylori-stratified analysis in East Asian ancestry individuals. 14
Supplementary Figure 16. Effect size comparison of distinct signals for DU and GU in East Asian ancestry individuals. 15
Supplementary Figure 17. Effect size comparison of distinct signals for DUonly and GUonly in BBJ1-180K. 16
Supplementary Figure 18. Effect size comparison of distinct signals for DU and GU in European ancestry individuals. 16
Supplementary Figure 19. Cross-cohort effect size comparison of distinct signals for DU and GU in East Asian ancestry individuals. 17
Supplementary Figure 20. Polygenicity estimation in East Asians using SBayesS. 18
Supplementary Figure 21. Effect size comparison of lead variants and secondary signals between PUD and GC in East Asian ancestry individuals. 18
Supplementary Figure 22. Cell-type specificity analysis in East Asian ancestry individuals using LDSC. 19
Supplementary Figure 23. Cell-type specificity analysis in East Asian ancestry individuals using MAGMA. 19
Supplementary Figure 24. Cell-type specificity analysis in European ancestry individuals using LDSC. 20
Supplementary Figure 25. Cell-type specificity analysis in European ancestry individuals using MAGMA. 20
Supplementary Figure 26. Cross-ancestry meta-analysis of cell-type specificity using LDSC. 21
Supplementary Figure 27. Cross-ancestry meta-analysis of cell-type specificity using MAGMA. 22

#### Abbreviations

PUD: peptic ulcer diseases
GU: gastric ulcers (including overlaps with duodenal ulcers)
GUonly: gastric ulcers only (excluding overlaps with duodenal ulcers)
DU: duodenal ulcers (including overlaps with gastric ulcers)
DUonly: duodenal ulcers only (excluding overlaps with duodenal ulcers)
BU: comorbidity of gastric ulcers and duodenal ulcers
GC: gastric cancers
HP: Helicobacter pylori
BBJ1: Biobank Japan 1st cohort
BBJ2: Biobank Japan 2nd cohort
BBJ1-180K: 180,000 individuals from BBJ1 (the main BBJ1 dataset)
BBJ1-12K: additional 12,000 individuals from BBJ1 (not included in the main dataset)
BBJ2-42K: 42,000 individuals from BBJ2
TMM-50K: 50,000 individuals from Tohoku Medical Megabank Project
UKB: UK Biobank
QC: quality control
GWAS: genome-wide association analysis
PheWAS: phenome-wide association analysis
Q-Q : quantile-quantile
$\lambda_{gc}$: genomic inflation factor
eQTL: expression quantitative trait loci
pQTL: protein quantitative trait loci
ATC: Anatomical Therapeutic Chemical code
EAS: East Asian
EUR: European
LDSC: linkage disequilibrium score regression

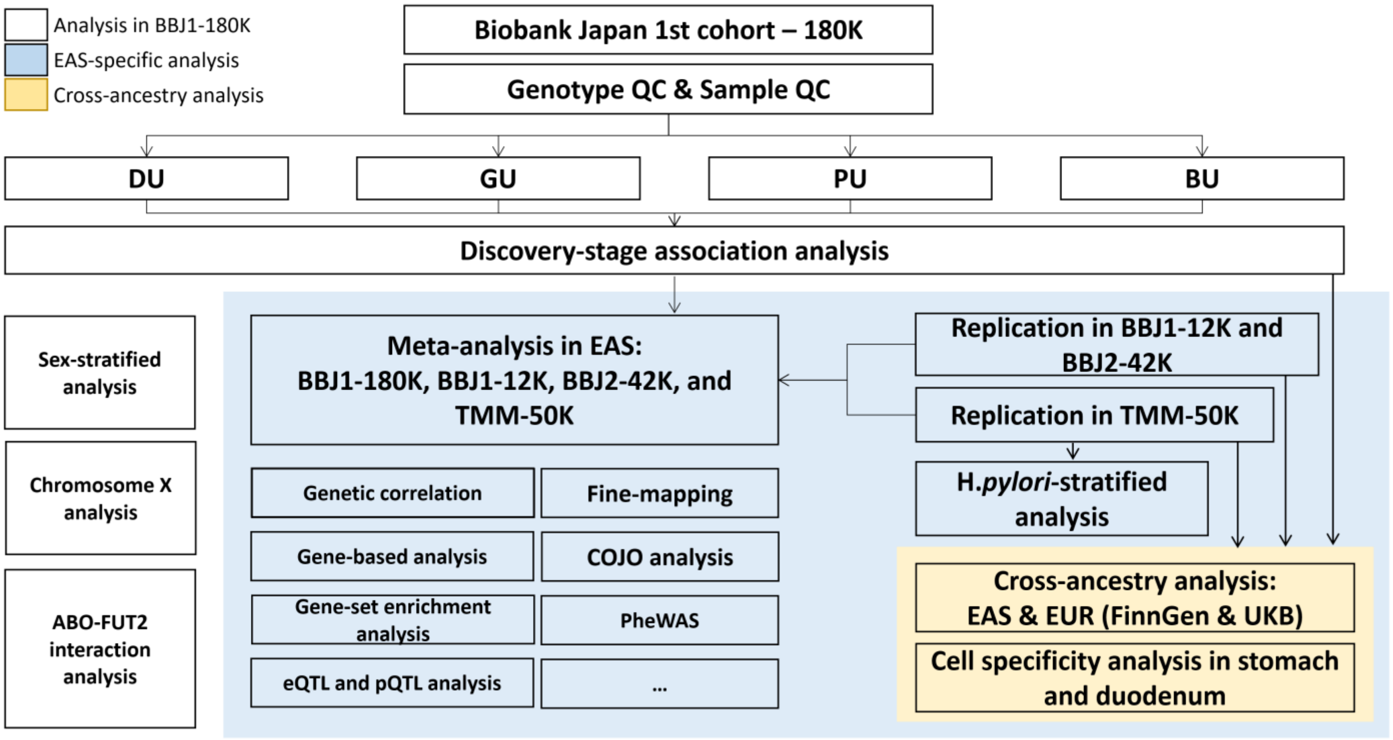


**Supplementary Figure 1. Study workflow.**

The three-stage design of this study was shown. We first performed discovery-stage GWAS for PUD and PUD subtypes in BBJ1-180K and then conducted replication in three independent studies. Next, East Asian-specific meta-analyses were conducted for PUD and PUD subtypes combining the four studies, and post-GWAS analyses were performed mainly using East Asian-specific summary statistics. Finally, we carried out cross-ancestry meta-analyses for PUD, DU, and GU, combining the four East Asian studies and GWAS in FinnGen and UKB.

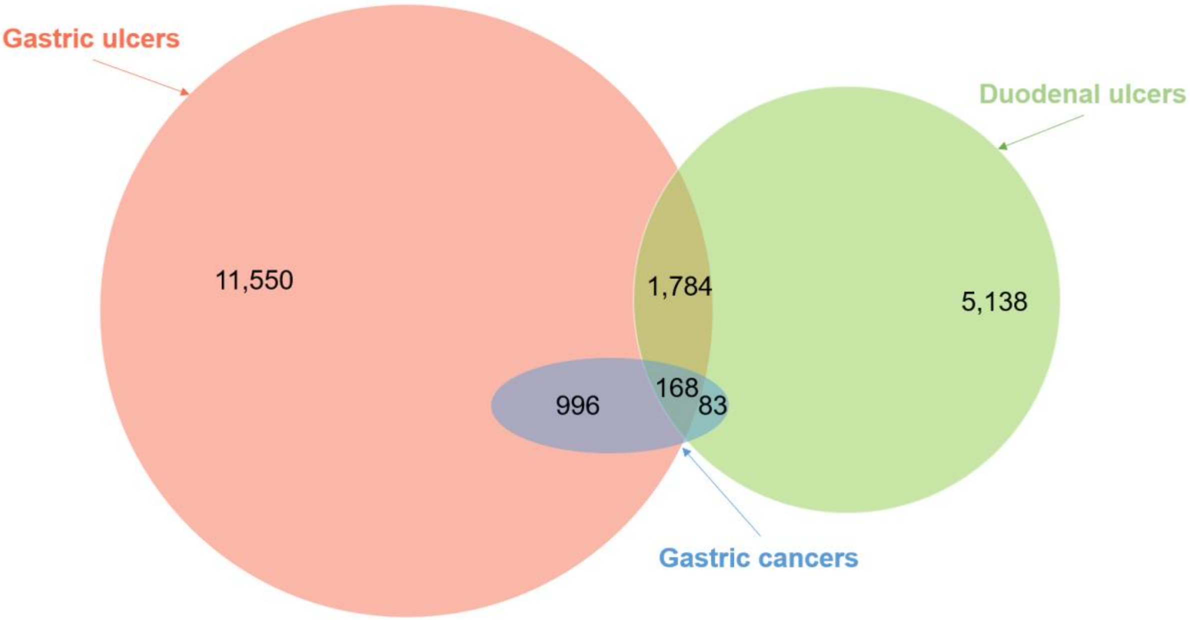


**Supplementary Figure 2. Venn diagram of cases used in the discovery-stage GWAS.**

Phenotype overlap among individuals with gastric ulcers, duodenal ulcers, and gastric cancers was shown for PUD cases in BBJ1-180K.

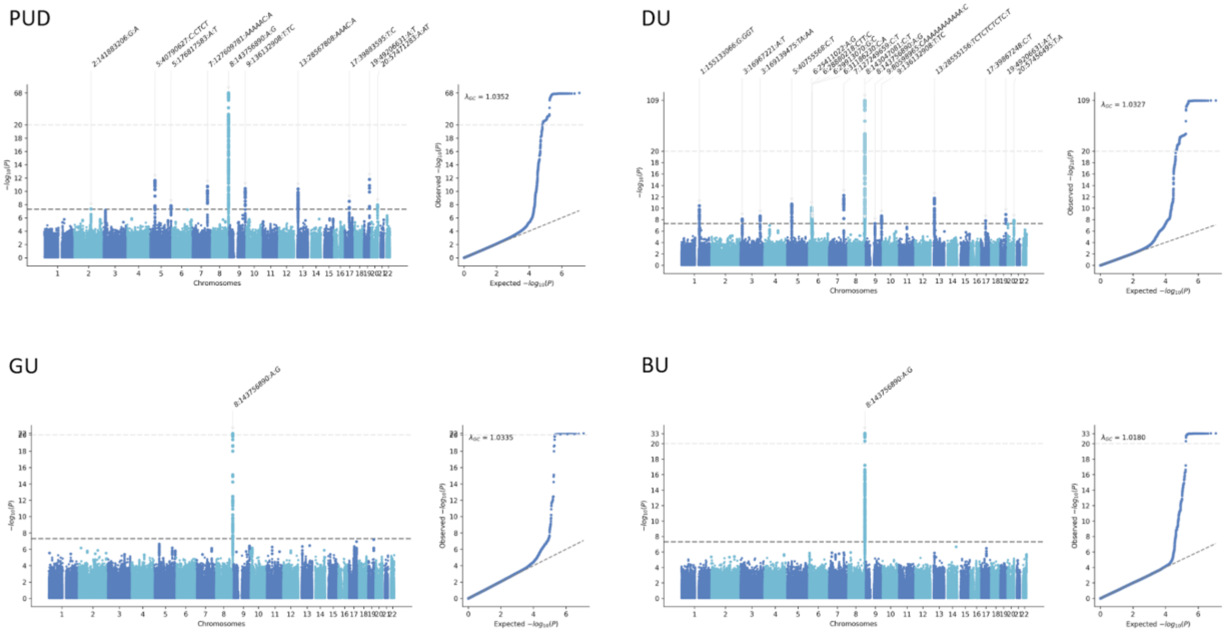


**Supplementary Figure 3. Manhattan plot and Q-Q plot for PUD and PUD subtypes from the discovery stage GWAS in BBJ1-180K.**

P values were derived from the discovery stage GWAS in BBJ1-180K. Genomic coordinates were mapped against GRCh37 (hg19). For variants above the top light grey dashed line($–\log_{10}$(P) >20 ), P values were rescaled. Significant loci were annotated with the lead variant. The bottom dark grey dashed line indicates the genome-wide significance threshold ($P<5 \times 10^{-8}$).

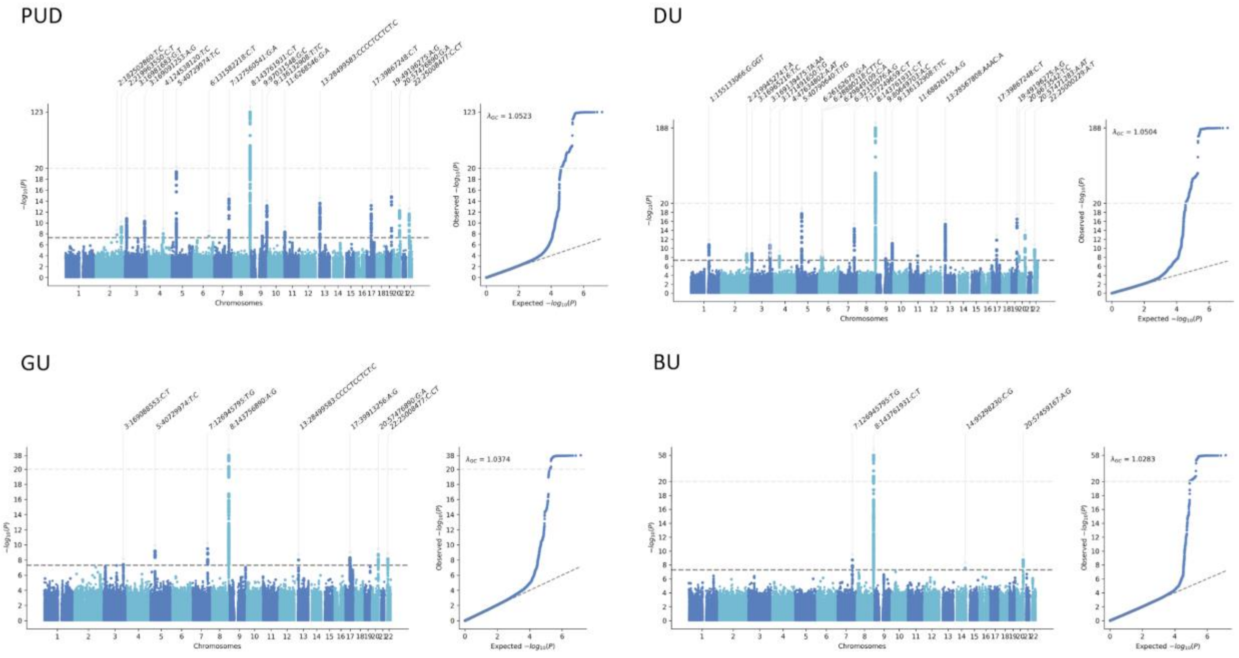


**Supplementary Figure 4. Manhattan plot and Q-Q plot for PUD and PUD subtypes from the East Asian-specific meta-analysis.**

P values were derived from the East Asian-specific meta-analyses combining the four studies. Genomic coordinates were mapped against GRCh37 (hg19). For variants above the top light grey dashed line($-\log_{10}(P) > 20$ ), P values were rescaled. Significant loci were annotated with the lead variant. The bottom dark grey dashed line indicates the genome-wide significance threshold ($P<5 \times 10^{-8}$).

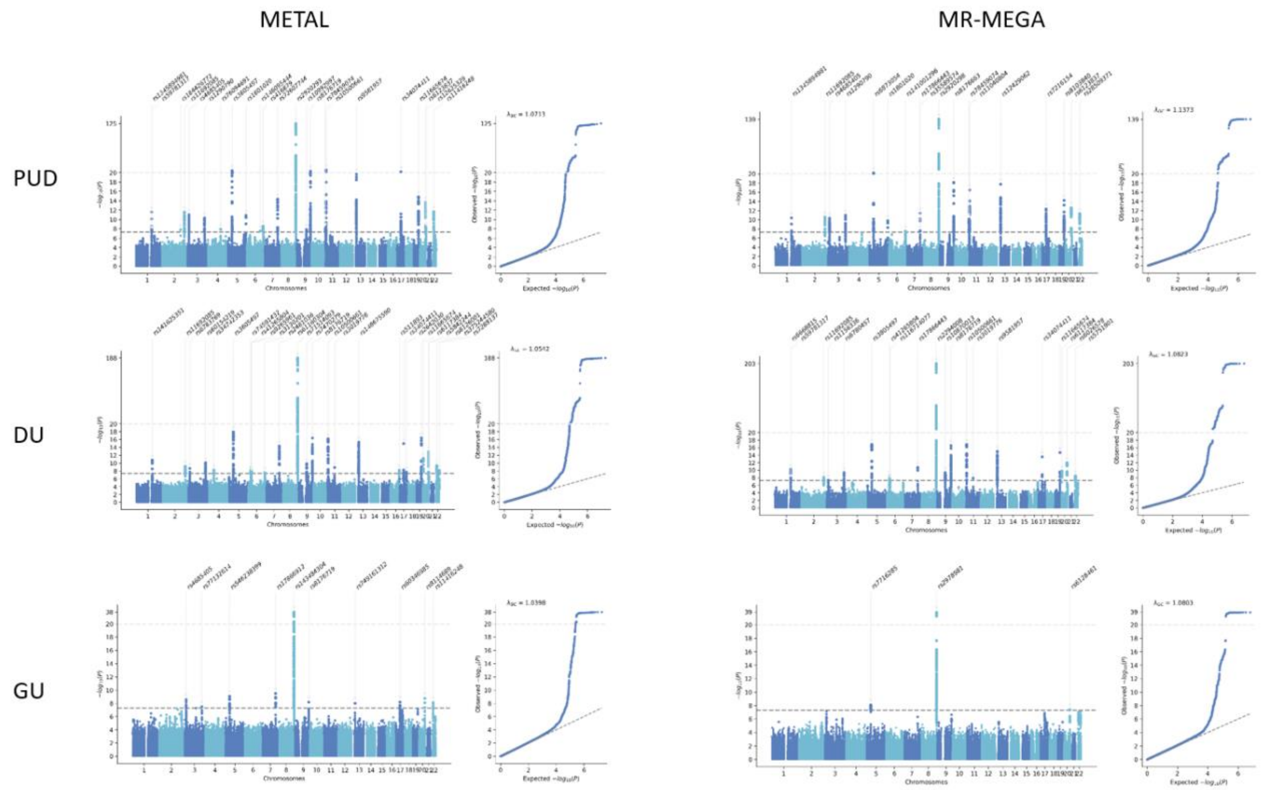


**Supplementary Figure 5. Manhattan plot and Q-Q plot for PUD and PUD subtypes from the cross-ancestry meta-analysis.**

P values were derived from the cross-ancestry meta-analyses using either METAL or MR-MEGA. For summary statistics from MR-MEGA, P values were recalculated from Chi-square statistics. Genomic coordinates were mapped against GRCh37 (hg19). For variants above the top light grey dashed line($-\log_{10}(P) > 20$ ), P values were rescaled. Significant loci were annotated with the lead variant. The bottom dark grey dashed line indicates the genome-wide significance threshold ($P<5 \times 10^{-8}$).

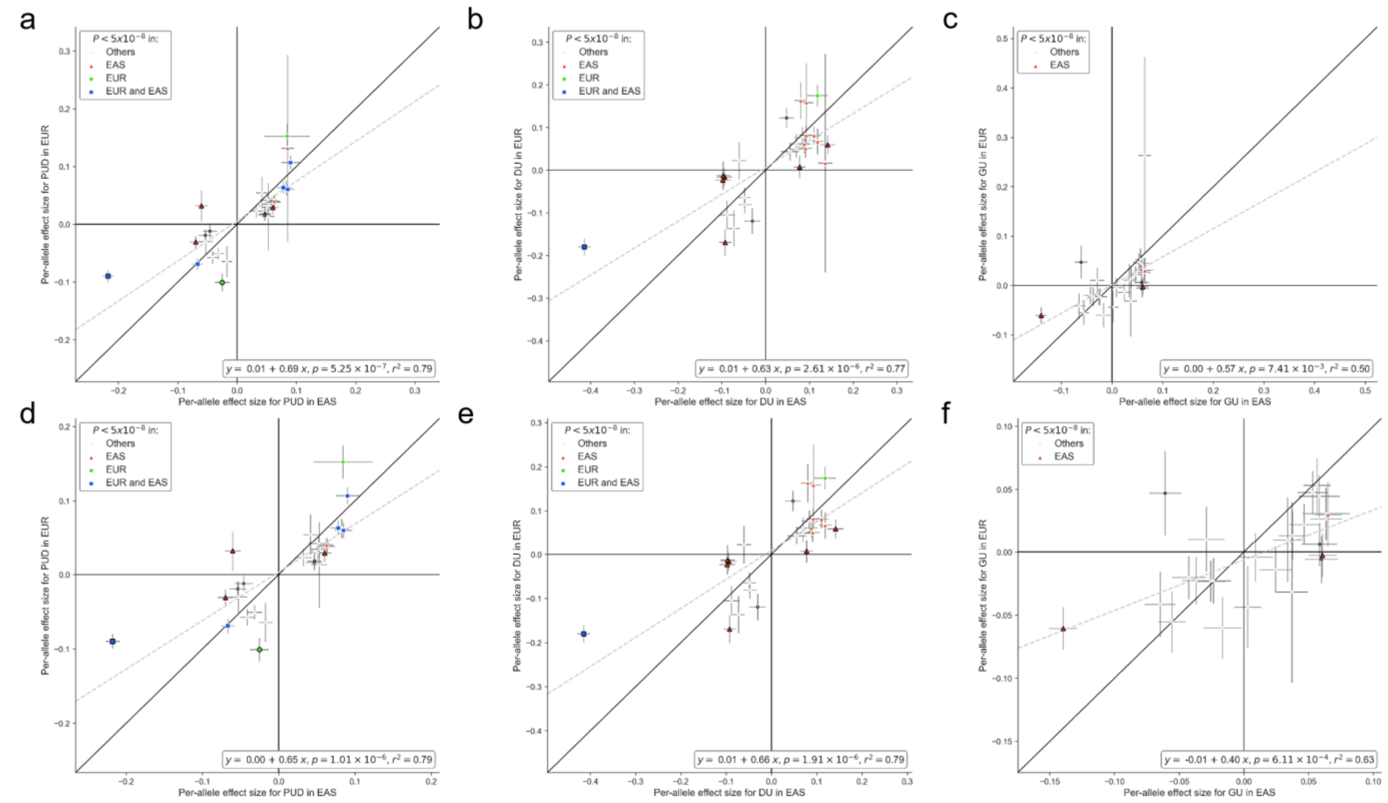


**Supplementary Figure 6. Cross-ancestry effect size comparison of lead variants for PUD and PUD subtypes.**

Per-allele effect size(β) comparison using EAS-specific and EUR-specific summary statistics for PUD and PUD subtypes. Lead variants associated with PUD or any subtypes in EAS-specific, EUR-specific, or cross-ancestry meta-analysis were selected for comparison. The most significant associations were shown if overlapping variants exist (interval < 500 kb). Variants with significant heterogeneity ($P_{het} < 0.05$) were denoted by the black marker edges. The grey dashed line represents the fitted linear regression line. **a**.28 available variants (existing in both datasets). **b**. 27 available variants. **c**. 27 available variants. **d**.27 available variants with MAF>0.01. **e**. 26 available variants with MAF>0.01. **f**. 26 available variants with MAF>0.01.

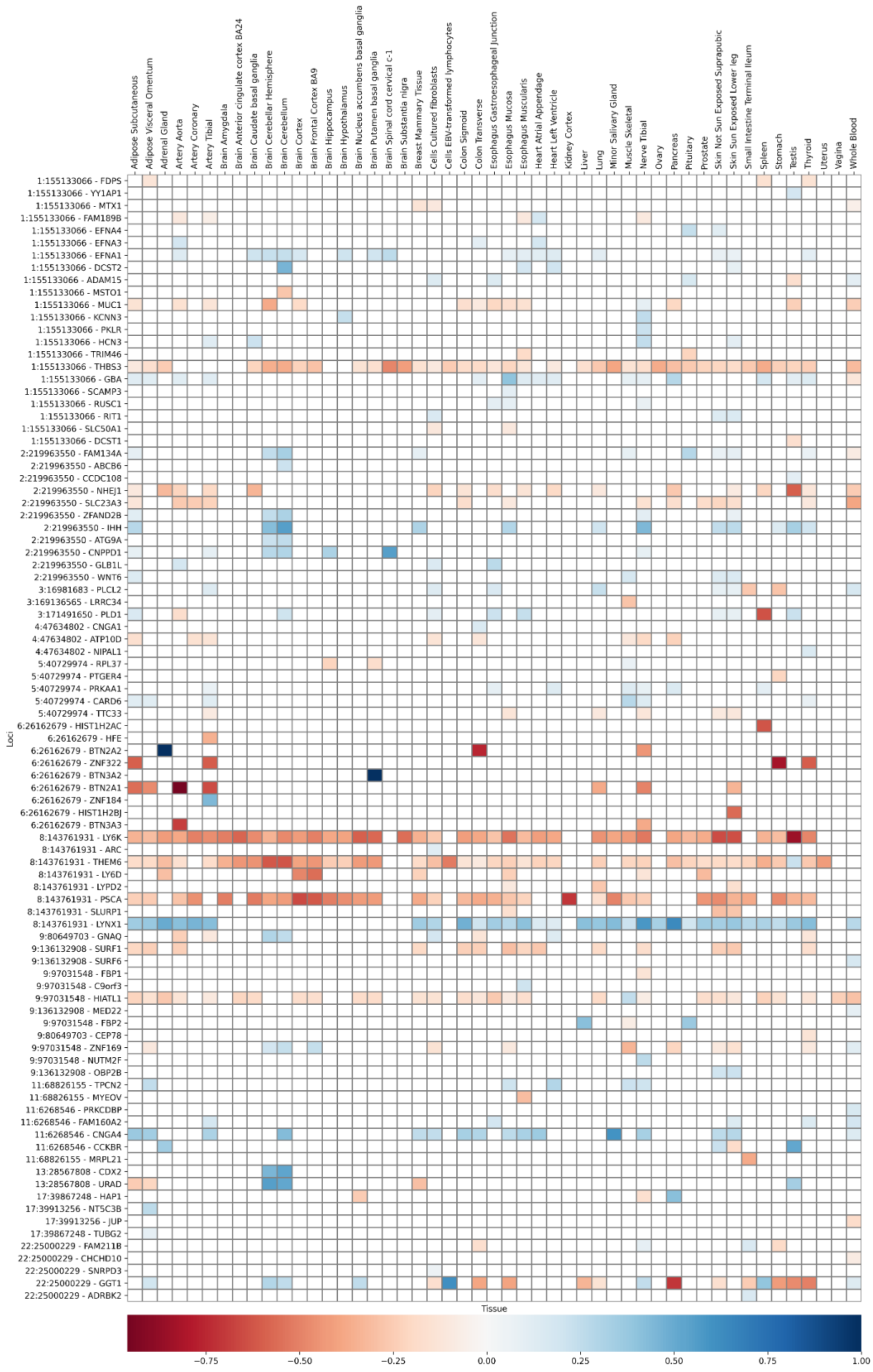

**Supplementary Figure 7. Overlap between PUD signals and significant cis-eQTL variants of the GTEx database.**

Overlap of significant cis-eQTL variants(FDR < 0.05) of the GTEx version 8 datasets[9] with lead variants in novel loci or its LD proxy ($r^2$>0.6 in 1KG EAS or EUR populations[10]) in 41 tissue types. Square colors represent the normalized beta values of the eQTL allele that is in LD with the PUD risk allele. For each transcript, only the most significant eQTL association was shown. Columns represent the tissue types, and rows show the genome coordinates of PUD lead variants (NCBI Build 37) and target transcripts of the overlapping eQTL association.

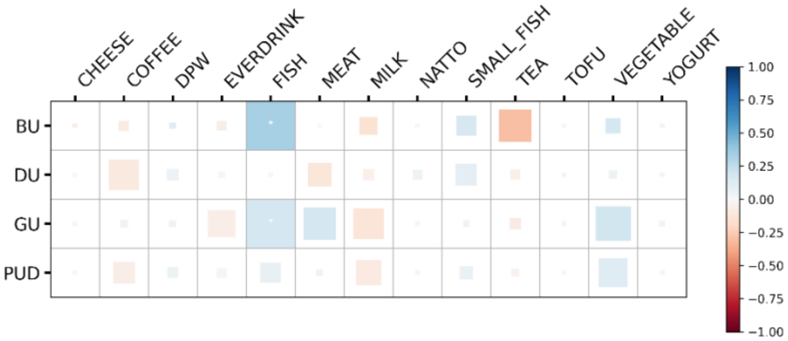


**Supplementary Figure 8. Genetic correlation heatmap of PUD and dietary habits in East Asian population.**

Genetic correlation among PUD and dietary habits in BBJ1-180K[11](**Methods**). None of the genetic correlations reached the significance threshold (FDR<0.05) after corrections. The size of the colored square in each cell is proportional to the $-\log_{10}(P)$ value of its corresponding correlation. *, $P < 0.05$.

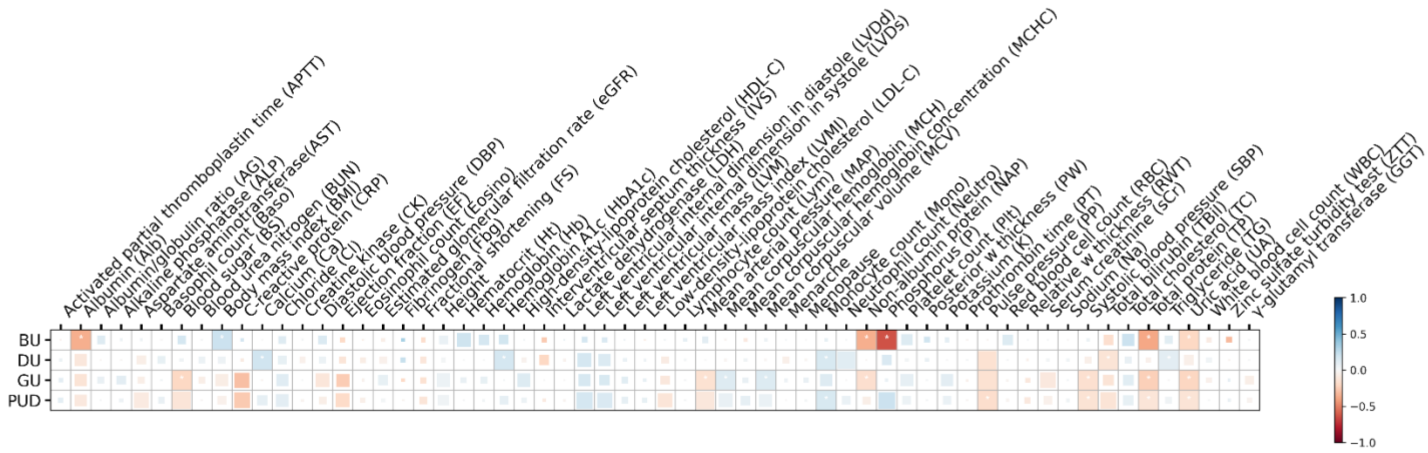


**Supplementary Figure 9. Genetic correlation heatmap of PUD and quantitative traits in East Asian population.**

Genetic correlation among PUD and quantitative traits in BBJ1-180K[12](**Methods**). None of the genetic correlations reached the significance threshold (FDR<0.05) after corrections. The size of the colored square in each cell is proportional to the $-\log_{10}(P)$ value of its corresponding correlation. *, $P < 0.05$.

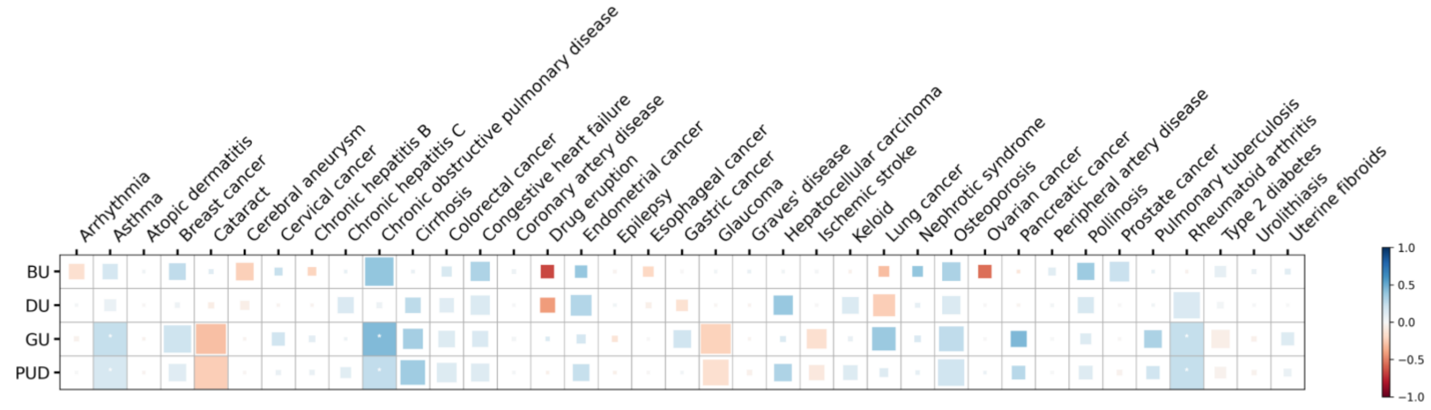


**Supplementary Figure 10. Genetic correlation heatmap of PUD and binary traits in East Asian population.**

Genetic correlation among PUD and binary traits in BBJ1-180K[13](**Methods**). None of the genetic correlations reached the significance threshold (FDR<0.05) after corrections. The size of the colored square in each cell is proportional to the $-\log_{10}(P)$ value of its corresponding correlation. *, $P < 0.05$.

.

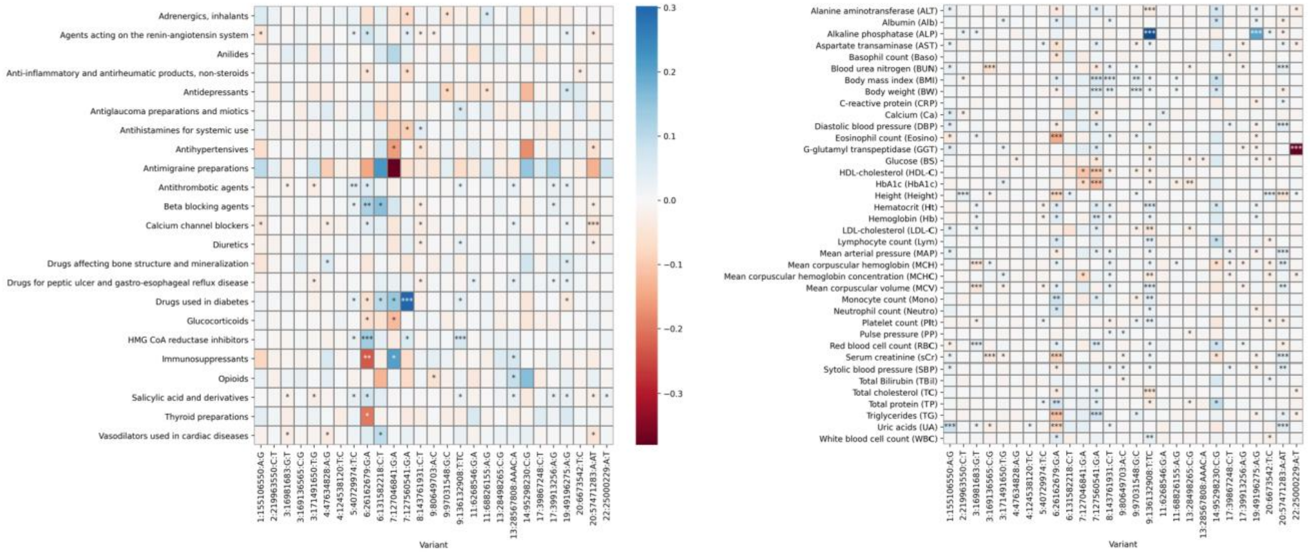


**Supplementary Figure 11. PheWAS heatmap of PUD risk variants with ATC codes and quantitative traits.**

Lead variants and independent secondary signals associated with PUD or any subtypes in the EAS population were selected for PheWAS lookup. a, ATC codes. b, quantitative traits. The most significant associations were selected if overlapping variants exist (interval < 500 kb). Summary statistics for ATC codes and quantitative traits were obtained from previous GWAS in BBJ1[14]. Per-allele effect sizes (log(OR)) of the PUD risk alleles were shown. *, P < 0.05 (Nominal significance). **, $P < 8.6 \times 10^{-6}$ (Bonferroni correction). ***, $P < 5.0 \times 10^{-8}$ (Genome-wide significance).

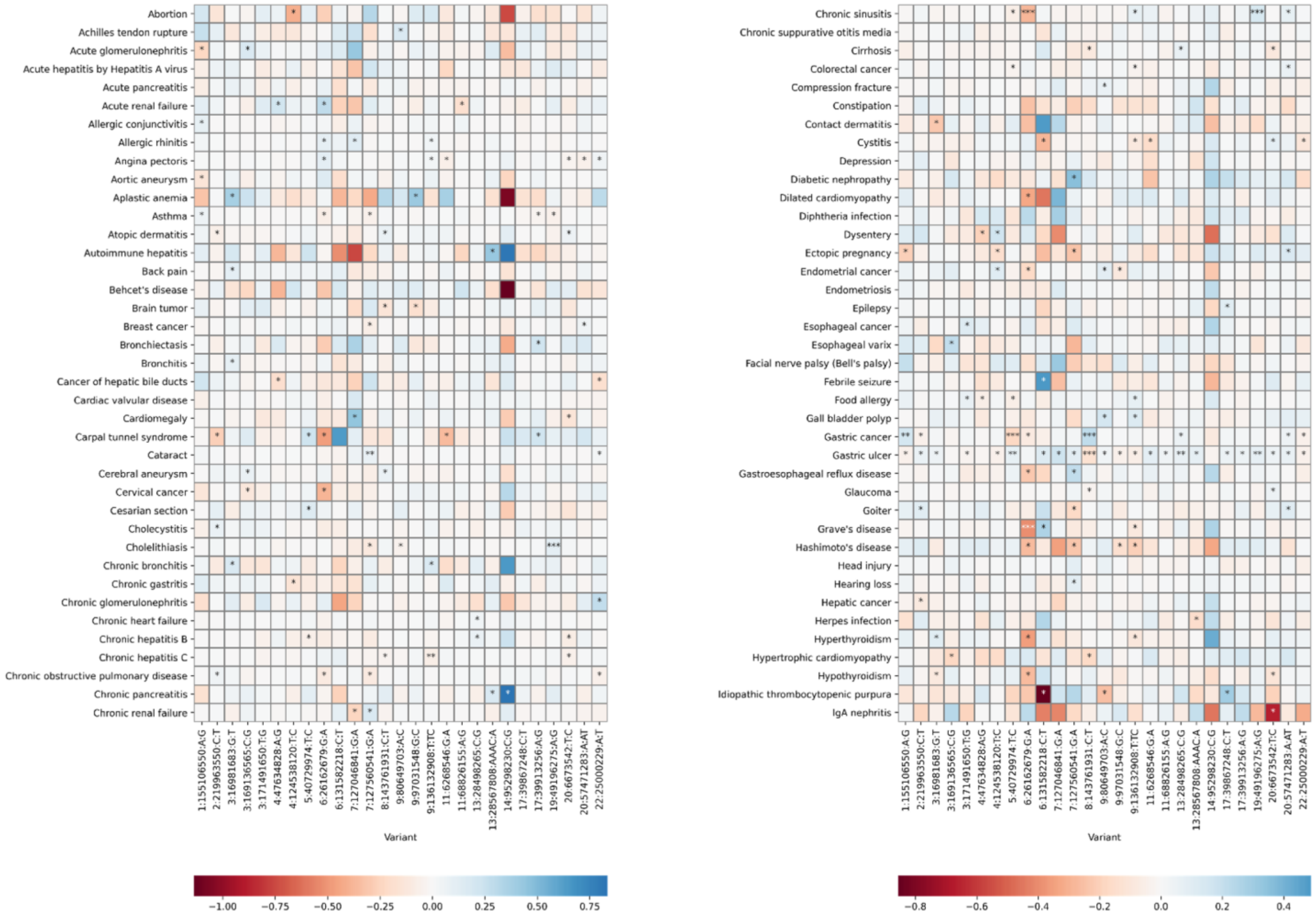


**Supplementary Figure 12. PheWAS heatmap of PUD risk variants with binary traits (Part 1/2).**

Lead variants and independent secondary signals associated with PUD or any subtypes in the EAS population were selected for PheWAS lookup. The most significant associations were selected if overlapping variants exist (interval < 500 kb). Summary statistics for binary traits were obtained from previous GWAS in BBJ1-180K[14]. Per-allele effect sizes (log(OR)) of the PUD risk alleles were shown. *, P < 0.05 (Nominal significance). **, $P < 8.6 \times 10^{-6}$ (Bonferroni correction). ***, $P < 5.0 \times 10^{-8}$ (Genome-wide significance).

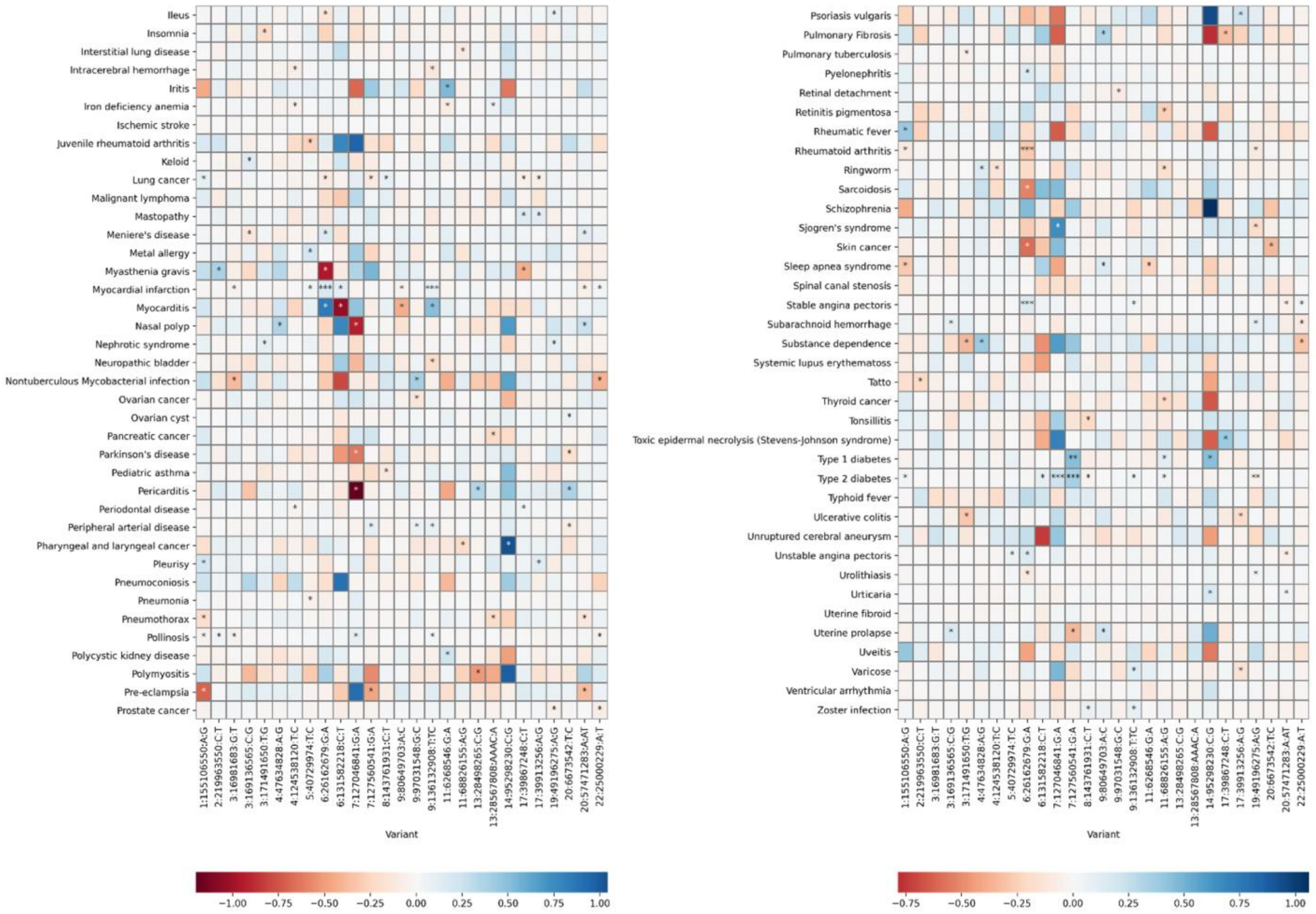


**Supplementary Figure 13. PheWAS heatmap of PUD risk variants with binary traits (Part 2/2).**

Lead variants and independent secondary signals associated with PUD or any subtypes in the EAS population were selected for PheWAS lookup. The most significant associations were selected if overlapping variants exist (interval < 500 kb). Summary statistics for binary traits were obtained from previous GWAS in BBJ1-180K[14]. Per-allele effect sizes (log(OR)) of the PUD risk alleles were shown. *, P < 0.05 (Nominal significance). **, $P < 8.6 \times 10^{-6}$ (Bonferroni correction). ***, $P < 5.0 \times 10^{-8}$ (Genome-wide significance).

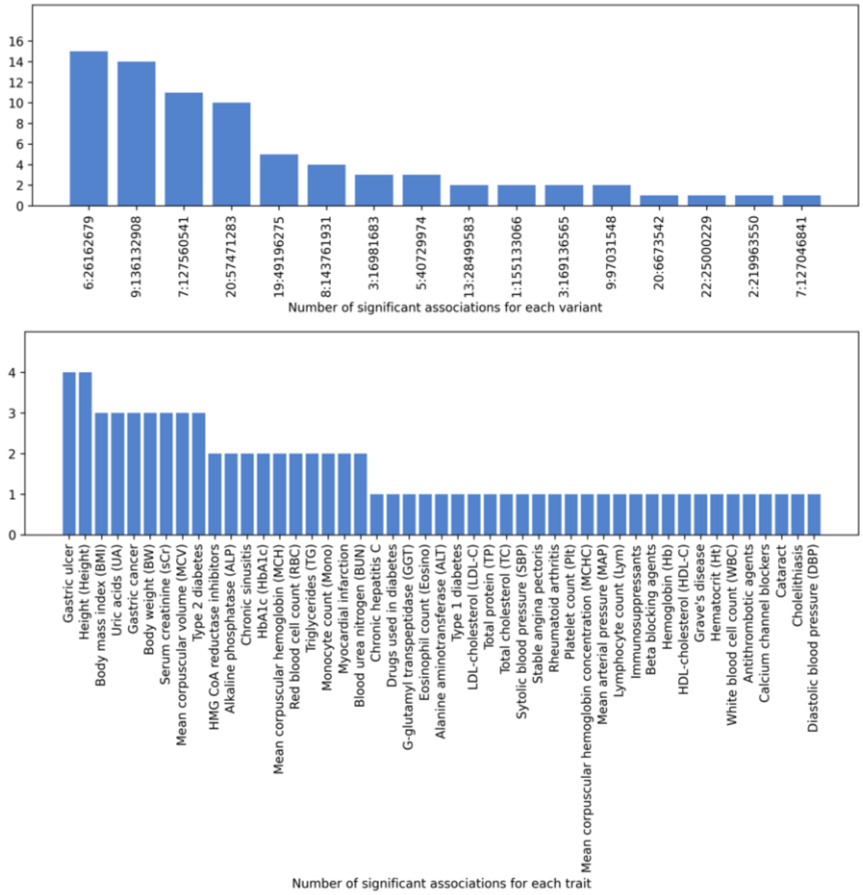


**Supplementary Figure 14. Summary of significant associations identified in PheWAS.**

Genome-wide significant associations identified in PheWAS lookup were summarized for each variant (top) or each trait (bottom).

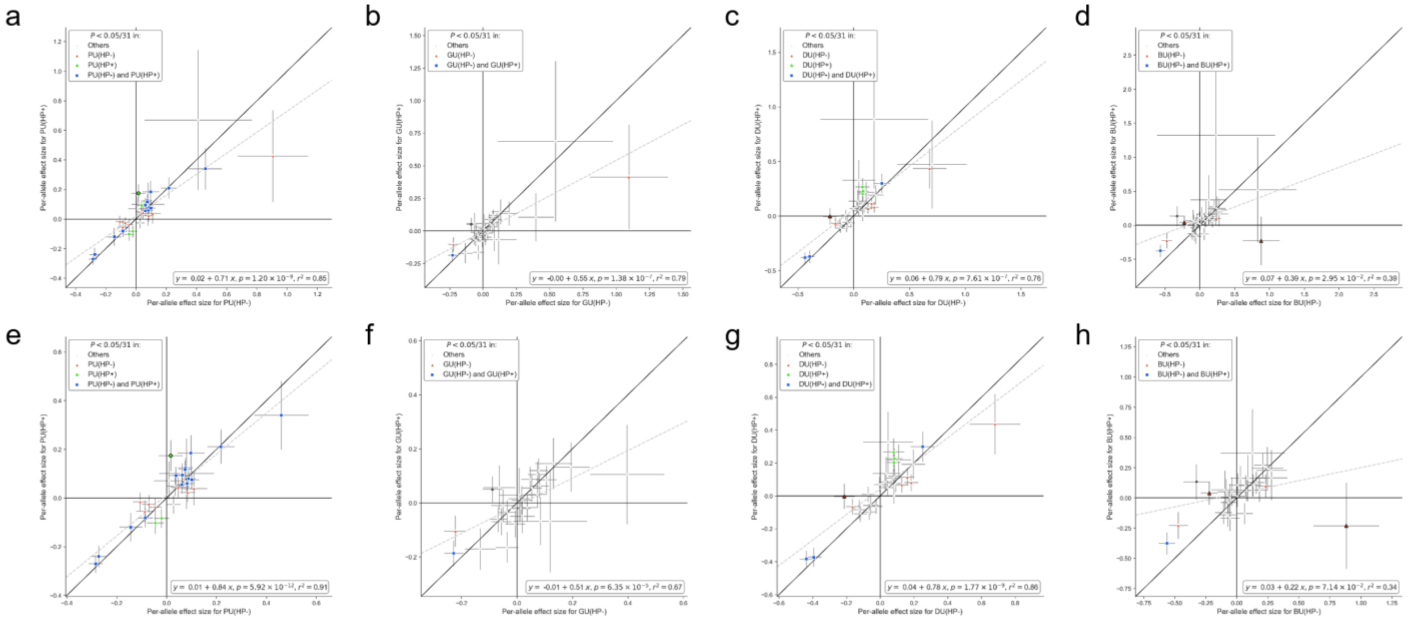

**Supplementary Figure 15. Effect size comparison of lead variants and secondary signals for H.pylori-stratified analysis in East Asian ancestry individuals.**

Per-allele effect size(β) comparison using summary statistics from H.pylori-stratified analysis. PUD(HP+), H.pylori-positive PUD; PUD(HP-), H.pylori-negative PUD; GU(HP+), H.pylori-positive GU; GU (HP-), H.pylori-negative GU; DU(HP+), H.pylori-positive DU; DU (HP-), H.pylori-negative DU; BU(HP+), H.pylori-positive BU; BU (HP-), H.pylori-negative BU. Lead variants and independent secondary signals associated with PUD or any subtypes in the EAS population were selected for comparison. The most significant associations were shown if overlapping variants exist (interval < 500 kb). **a,e**, PUD. **b,f,** GU. **e,g**, DU. **d,h**, BU. **a,b,c,d**. 31 available variants (existing in both datasets) were shown. **e,f,g,h**. 29 available variants with MAF>0.01 were shown.

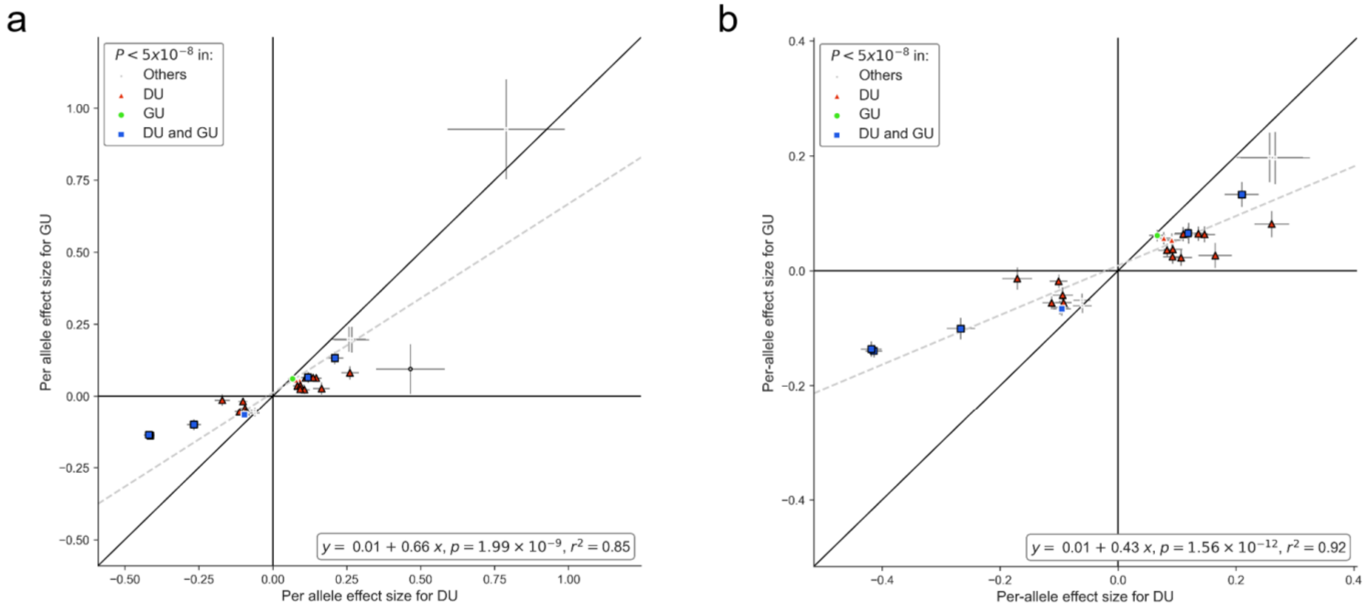


**Supplementary Figure 16. Effect size comparison of distinct signals for DU and GU in East Asian ancestry individuals.**

Per-allele effect size ( log(OR) ) comparison using EAS-specific summary statistics for DU and GU. Lead variants and independent secondary signals associated with PUD or any subtypes in the EAS population were selected for comparison. The most significant associations were shown if overlapping variants exist (interval < 500 kb). Variants with nominal significant heterogeneity ($P_{het} < 0.05$) were denoted by black marker edges. The grey dashed line represents the fitted linear regression line, whose parameters were shown in the bottom right; p, p-value obtained from t-test for slope. Marker colors denote the GWAS in which the significant variants were identified; Others, PUD or BU. **a.** all 31 available variants (existing in both datasets) were shown. **b.** 29 variants with MAF>0.01 were shown.

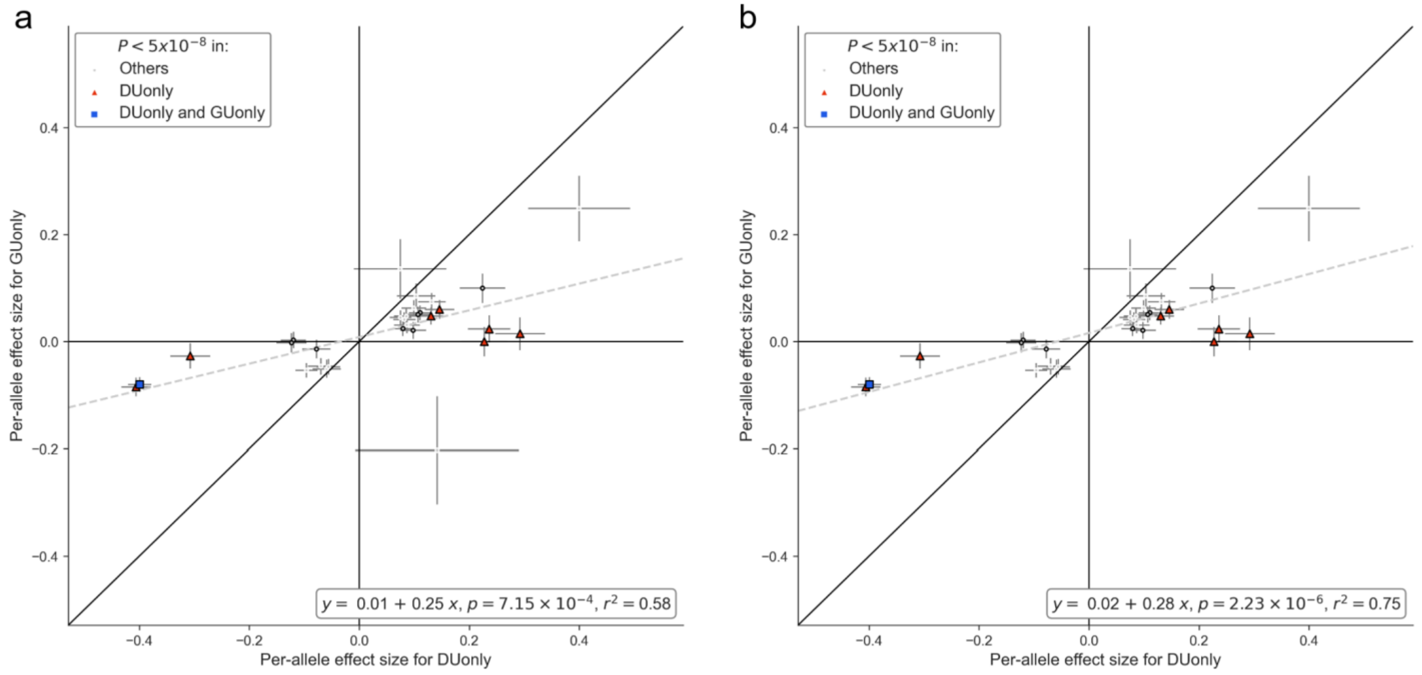


**Supplementary Figure 17. Effect size comparison of distinct signals for DUonly and GUonly in BBJ1-180K.**

Per-allele effect size ( log(OR) ) comparison using summary statistics for DUonly and GUonly in BBJ1-180K. Lead variants and independent secondary signals associated with PUD or any subtypes in the EAS population were selected for comparison. The most significant associations were shown if overlapping variants exist (interval < 500 kb). Variants with significant heterogeneity ($P_{het} < 0.05$) were denoted by black marker edges. The grey dashed line represents the fitted linear regression line, whose parameters were shown in the bottom right; p, p-value obtained from t-test for slope. Marker colors denote the GWAS in which the significant variants were identified; Others, PUD or BU. **a.** 30 available variants (existing in both datasets) were shown. **b.** 29 available variants with MAF>0.01 were shown.

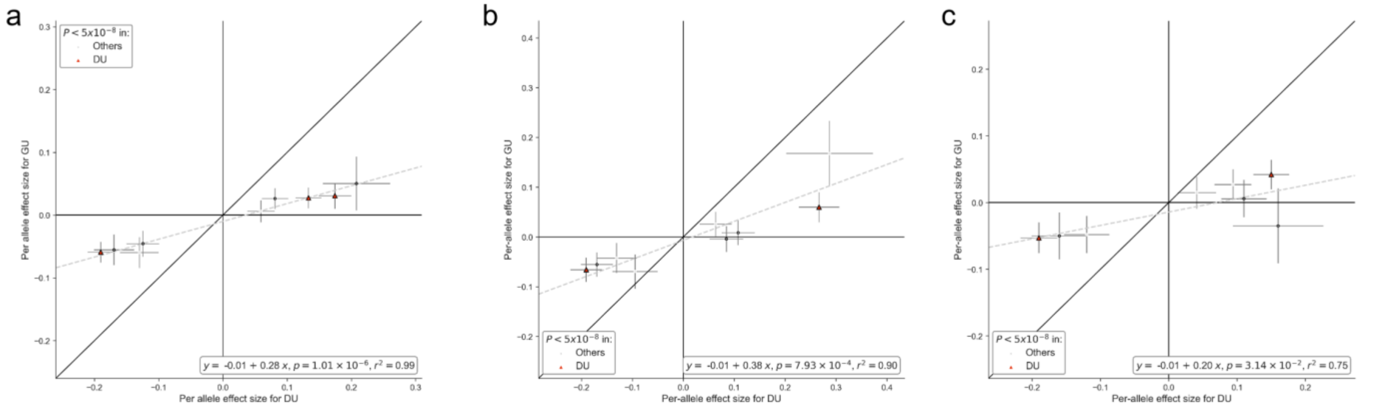


**Supplementary Figure 18. Effect size comparison of distinct signals for DU and GU in European ancestry individuals.**

Per-allele effect size ( log(OR) ) comparison using summary statistics for DU and GU derived from EUR-specific meta-analysis(**a**), GWAS in UKB(**b**), and GWAS in FinnGen(**c**). Details were described in **Methods**. Lead variants associated with PUD or any subtypes in the EUR population were selected for comparison. The most significant associations were shown if overlapping variants exist (interval < 500 kb). Variants with significant heterogeneity ($P_{het} < 0.05$) were denoted by black marker edges. The grey dashed line represents the fitted linear regression line, whose parameters were shown in the bottom right; p, p-value obtained from t-test for slope. Marker colors denote the GWAS in which the significant variants were identified; Others, PUD. **a,b.** 9 available variants (existing in both datasets) were shown. **c.** 8 available variants were shown.

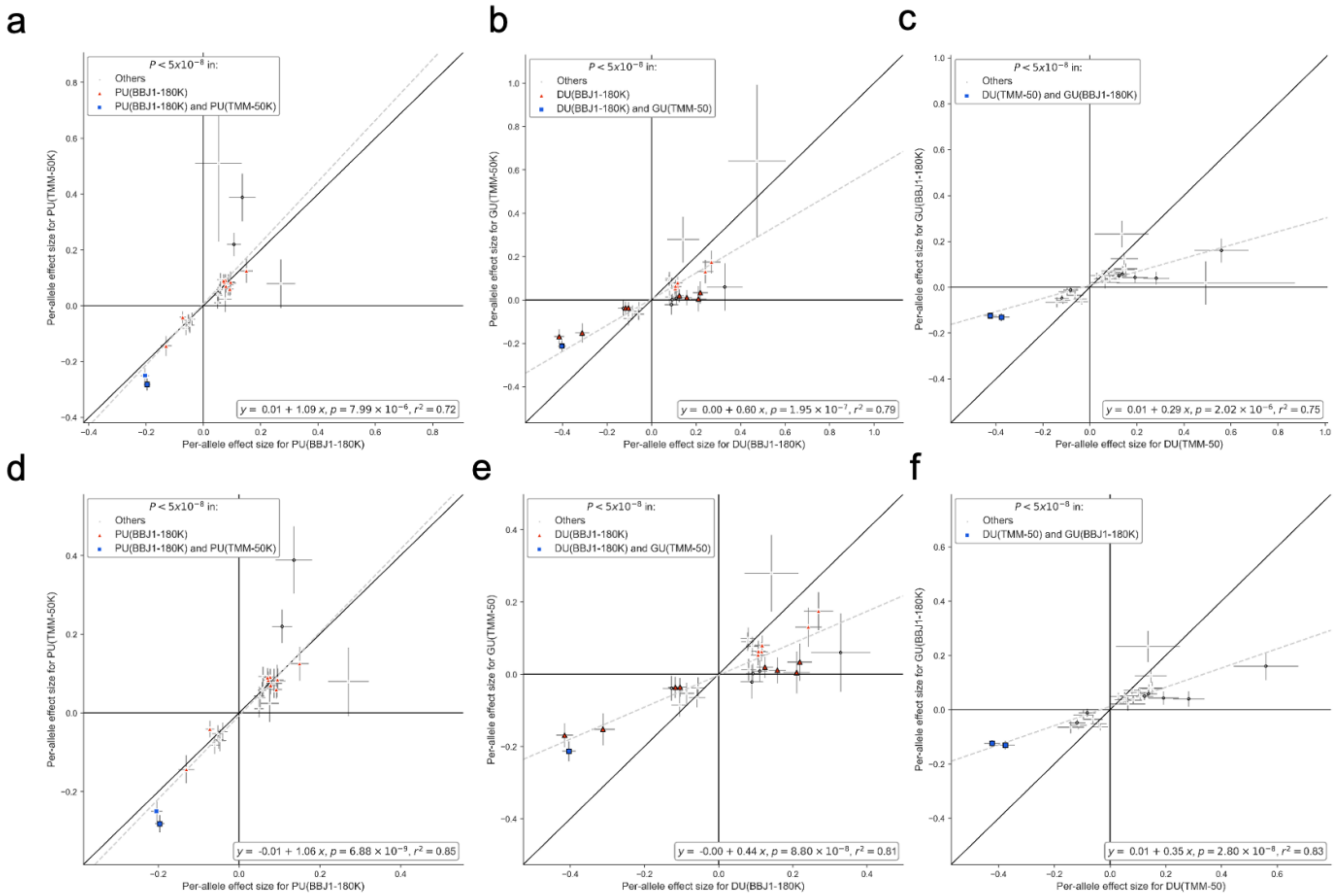


**Supplementary Figure 19. Cross-cohort effect size comparison of distinct signals for DU and GU in East Asian ancestry individuals.**

Per-allele effect size ( log(OR) ) comparison using summary statistics for PUD(**a,d**), DU(**b,e**), and GU(**c,f**) derived from GWAS in BBJ1-180K and TMM-50K. Lead variants and independent secondary signals associated with PUD or any subtypes in the EAS population were selected for comparison. The most significant associations were shown if overlapping variants exist (interval < 500 kb). For PUD, TMM-50K-derived statistics were compared with BBJ1-180K-derived statistics(**a,d**). For DU and GU, TMM-50K-derived GU statistics were compared with BBJ1-180K-derived DU statistics(**b,e**), and TMM-50K-derived DU statistics were compared with BBJ1-180K-derived GU statistics(**c,f**). Variants with significant heterogeneity ($P_{het} < 0.05$) were denoted by black marker edges. The grey dashed line represents the fitted linear regression line, whose parameters were shown in the bottom right; p, p-value obtained from t-test for slope. Marker colors denote the GWAS in which the significant variants were identified; Others, PUD. **a,** 28**, b,** 27**, c,** 27. available variants (existing in both datasets) were shown. **d,** 27**, e,** 26**, f,** 26, available variants with MAF>0.01 were shown.

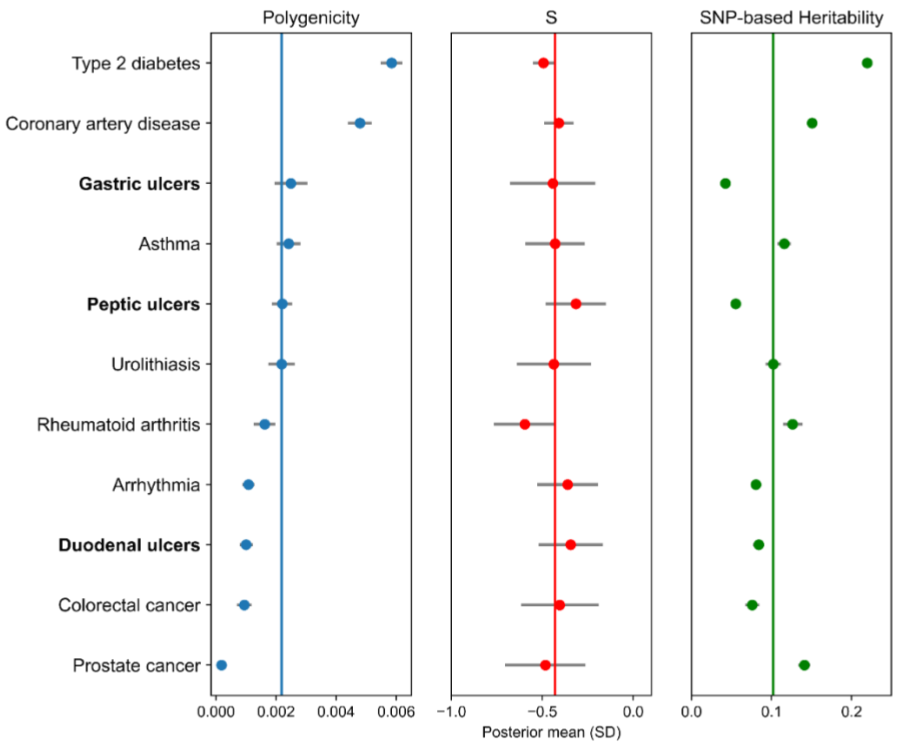


**Supplementary Figure 20. Polygenicity estimation in East Asians using SBayesS.**

Summary statistics for binary phenotypes from BBJ1[13], and meta-analysis of PUD and PUD subtypes in East Asians were used for SbayesS analysis. Only phenotypes that converged in the SbayesS model were shown. Phenotypes were ranked based on polygenicity estimates. SNP-based heritability estimates were on the liability scale. The vertical lines indicate the median estimates of the 11 phenotypes.

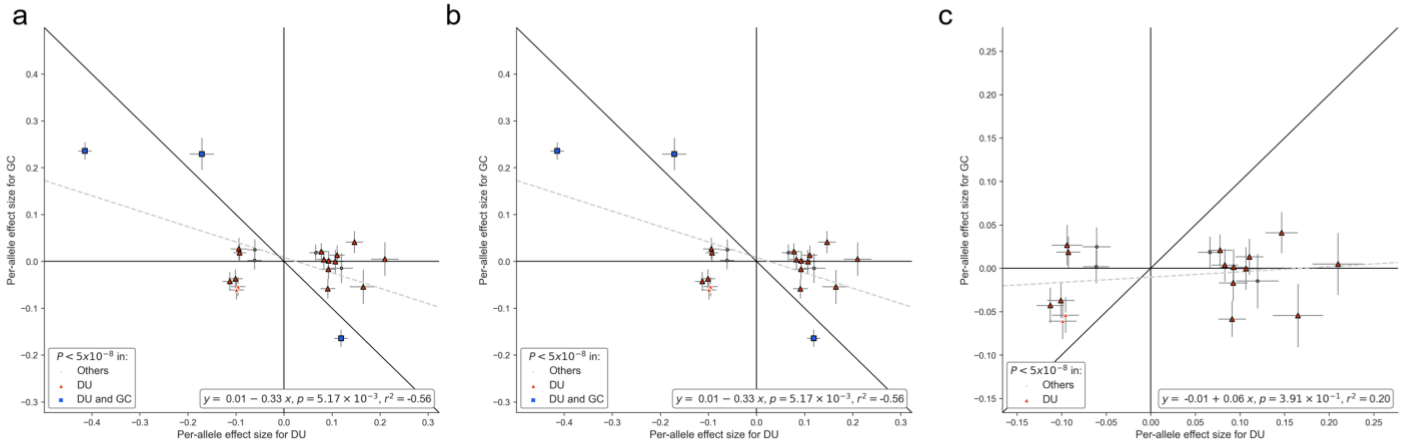


**Supplementary Figure 21. Effect size comparison of lead variants and secondary signals between PUD and GC in East Asian ancestry individuals.**

Per-allele effect size(β) comparison between DU and GC. Summary statistics for DU were obtained from EAS-specific meta-analysis. Summary statistics for GC were obtained from previous GWAS conducted in BBJ1 (Methods). Lead variants and independent secondary signals associated with PUD or any subtypes in the EAS population were selected for comparison. The most significant associations were shown if overlapping variants exist (interval < 500 kb). **a**. 23 available variants (existing in both datasets) were shown. **b**. 23 available variants with MAF>0.01 were shown. **c**. 20 variants (excluding lead variants at *EFNA1*, *PTGER4*, and *PSCA*) were shown.

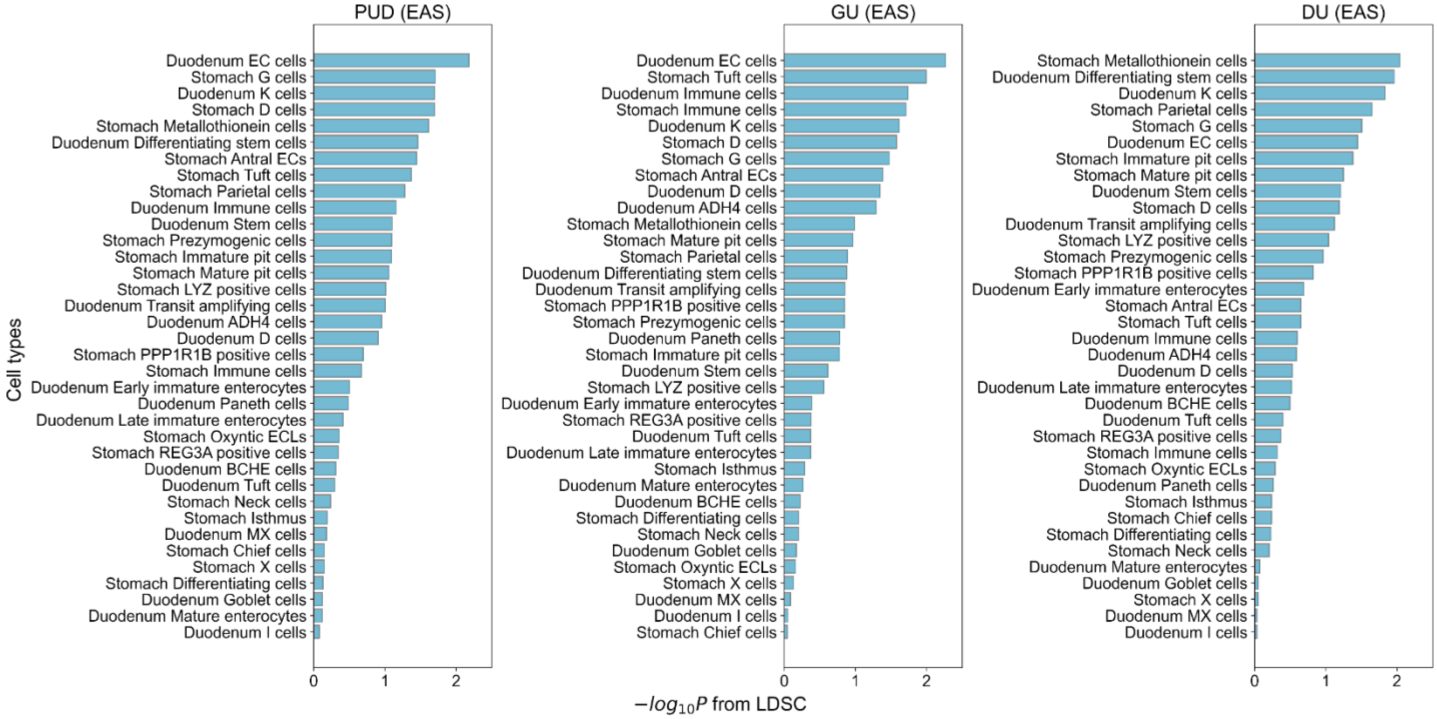


**Supplementary Figure 22. Cell-type specificity analysis in East Asian ancestry individuals using LDSC.**

Associations between PUD and cell types in the stomach and duodenum were analyzed using LDSC (testing for the enrichment of the 10% most specific genes in each cell type; **Methods**). East Asian-specific summary statistics were used in the analysis. X axis, –log10(P) derived from LDSC estimates. Color bars indicate whether the association is significant (Red, FDR<5%; Light blue, FDR>=5%).

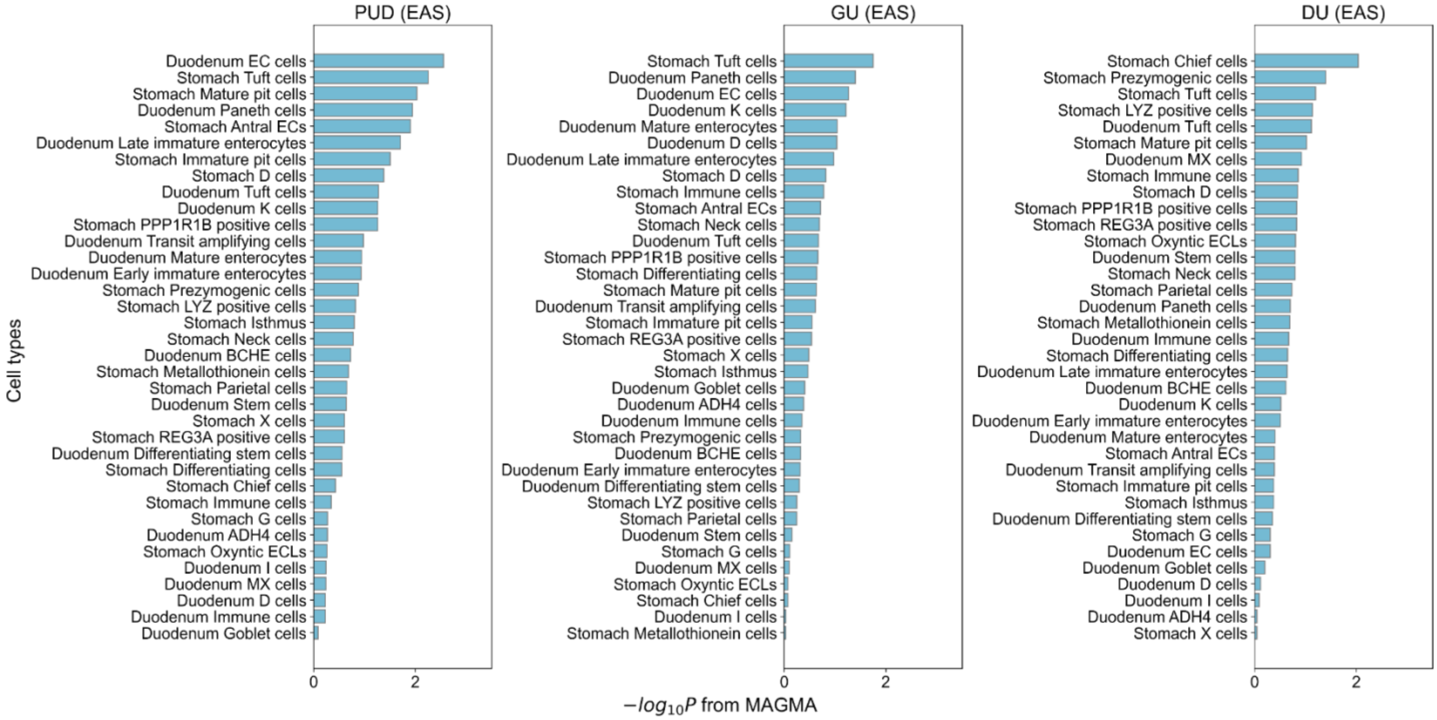


**Supplementary Figure 23. Cell-type specificity analysis in East Asian ancestry individuals using MAGMA.**

Associations between PUD and cell types in the stomach and duodenum were analyzed using MAGMA (testing for the enrichment of the 10% most specific genes in each cell type; **Methods**). East Asian-specific summary statistics were used in the analysis. X axis, –log10(P) derived from MAGMA estimates. Color bars indicate whether the association is significant (Red, FDR<5%; Light blue, FDR>=5%).

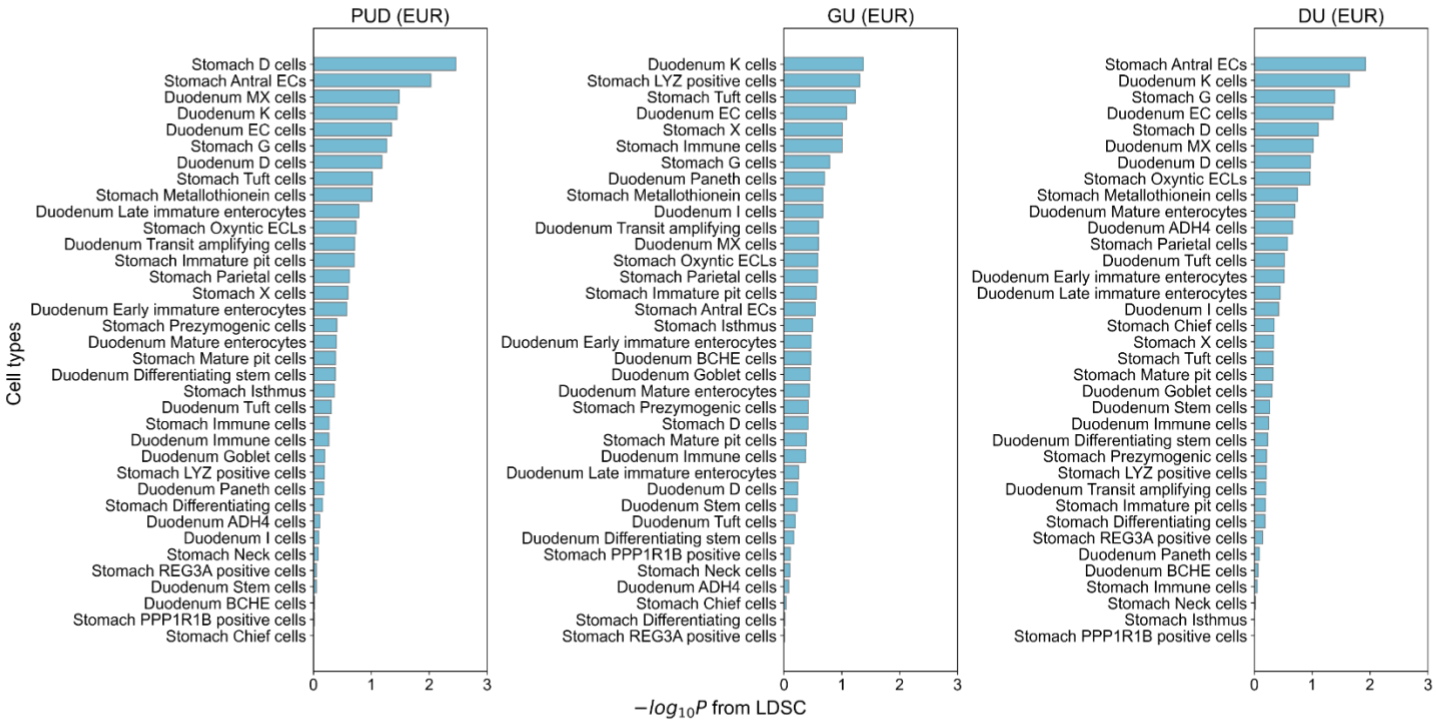


**Supplementary Figure 24. Cell-type specificity analysis in European ancestry individuals using LDSC.**

Associations between PUD and cell types in the stomach and duodenum were analyzed using LDSC (testing for the enrichment of the 10% most specific genes in each cell type; **Methods**). European-specific summary statistics were used in the analysis. X axis, –log10(P) derived from LDSC estimates. Color bars indicate whether the association is significant (Red, FDR<5%; Light blue, FDR>=5%).

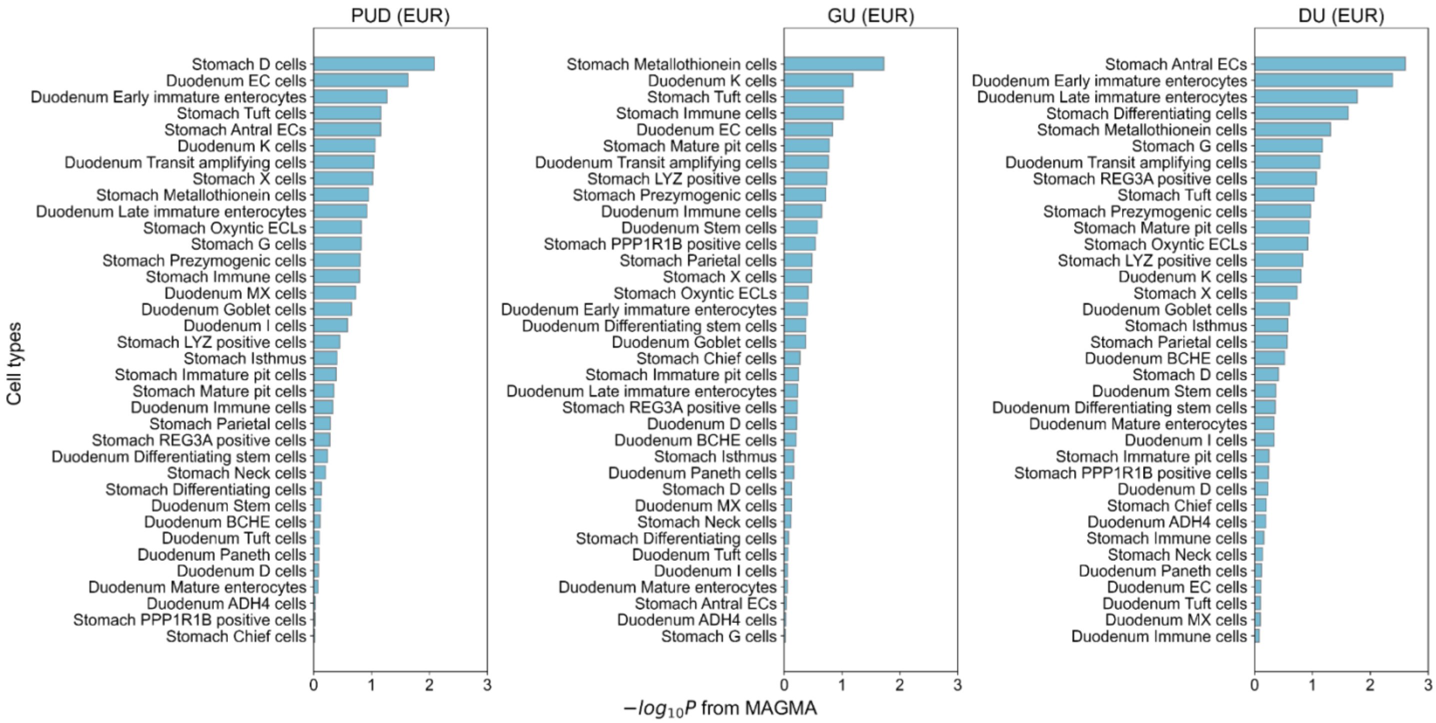


**Supplementary Figure 25. Cell-type specificity analysis in European ancestry individuals using MAGMA.**

Associations between PUD and cell types in the stomach and duodenum were analyzed using MAGMA (testing for the enrichment of the 10% most specific genes in each cell type; **Methods**). European-specific summary statistics were used in the analysis. X axis, –log10(P) derived from MAGMA estimates. Color bars indicate whether the association is significant (Red, FDR<5%; Light blue, FDR>=5%).

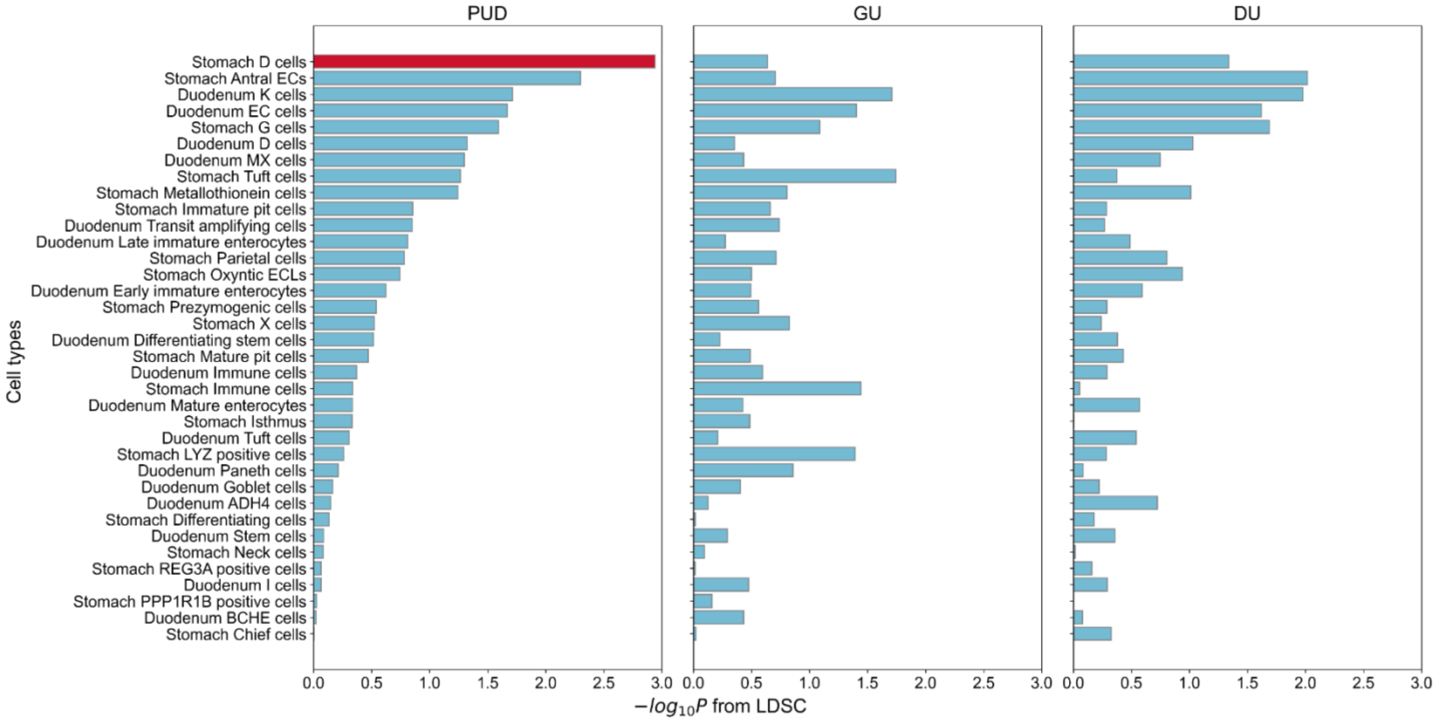


**Supplementary Figure 26. Cross-ancestry meta-analysis of cell-type specificity using LDSC.**

Associations between PUD and cell types in the stomach and duodenum were analyzed using LDSC (testing for the enrichment of the 10% most specific genes in each cell type). Inverse variance weighted meta-analysis was performed combining statistics from EAS and EUR ancestries. X axis, –log10(P) derived from meta-analyzed estimates. Color bars indicate whether the enrichment is significant (Red, FDR<5%; Light blue, FDR>=5%) in meta-analyzed results.

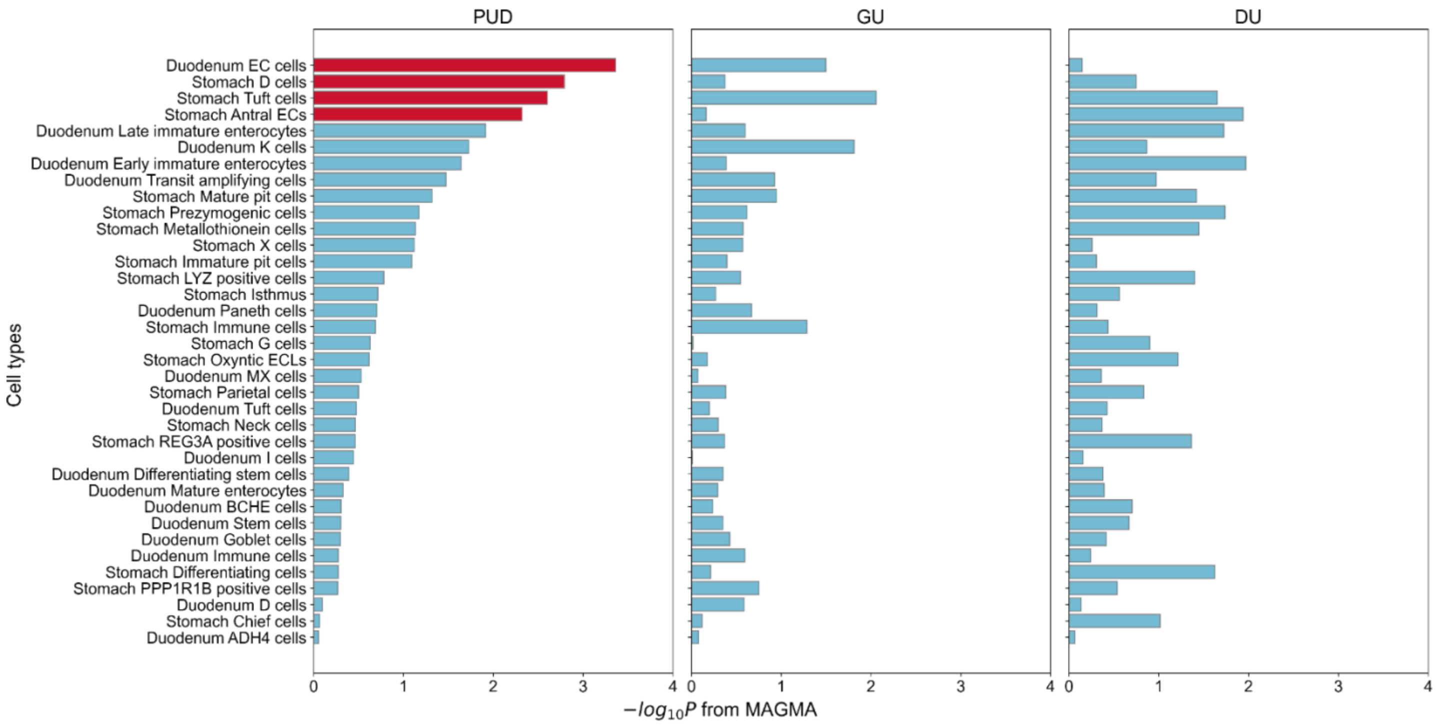


**Supplementary Figure 27. Cross-ancestry meta-analysis of cell-type specificity using MAGMA.**

Association between PUD and cell types in the stomach and duodenum were analyzed using MAGMA (testing for the enrichment of the 10% most specific genes in each cell type). Inverse variance weighted meta-analysis was performed combining statistics from EAS and EUR ancestries. X axis, –log10(P) derived from meta-analyzed estimates. Color bars indicate whether the enrichment is significant (Red, FDR<5%; Light blue, FDR>=5%) in meta-analyzed results.
